# Learning interventions and training methods in health emergencies: A scoping review

**DOI:** 10.1101/2023.08.08.23293718

**Authors:** Heini Utunen, Giselle Balaciano, Elham Arabi, Anna Tokar, Jane Noyes

## Abstract

**Background:** Keeping the health workforce and the public informed with the latest evolving knowledge is critical to preventing, detecting and responding to infectious disease outbreaks or other health emergencies. Having a well informed, ready, willing, and skilled workforce and an informed public can help save lives, reduce diseases and suffering, and minimize socio-economic loss in affected communities and countries. Providing support and opportunities for learning in health emergencies is much needed for capacity building. In this paper, ‘learning intervention’ refers to the provision of ad-hoc, focused, or personalized training sessions with the goal of preparing the health workers for emergencies or filling specific knowledge or skill gaps. We refer to ‘training methods’ as instructional design strategies used to teach someone the necessary knowledge and skills to perform a task.

**Methods:** We conducted a scoping review to map and better understand what learning interventions and training methods have been used in the different types of health emergencies and with whom. Studies were identified by searching Pubmed/Medline, Embase and consulting with experts.

**Results:** Of the 319 records that were included, contexts related to COVID-19, disasters in general, Ebola and wars were most frequently covered. Four topic areas were identified: Knowledge acquisition, Emergency Plans, Impact of the learning intervention, and training methods. Much of the evidence was based on observational methods with few trials, which likely reflects the unique context of each health emergency. Evolution of methods was apparent, particularly in virtual learning. Learning during health emergencies appeared to improve knowledge, management, quality of life, satisfaction and clinical outcomes.

**Conclusion:** This is the first scoping review to map the evidence, which serves as a first step to develop urgently needed global guidance to further improve the quality and reach of learning interventions and training methods in this context.

## Introduction

Learning in health emergencies can provide a foundation to build capacity for emergency preparedness and response specific to different health emergencies(1) (e.g., biological, environmental, armed conflicts, deliberate acts of terrorism, industrial accidents), especially in low and middle income countries where health systems need to be further strengthened(2). A ready, willing, and able workforce is required that can be called upon in health emergencies to help save lives, reduce diseases and suffering, and minimize socio-economic loss in affected communities and countries. In today’s interconnected landscape,an educated public is also needed to champion measures for a strong emergency preparedness and effective response(1). This can be achieved by guiding, standardizing and facilitating the delivery of life-saving knowledge, first to frontline workers in health emergencies and second to the public. Learning paves the way for strengthened health literacy and understanding of health communication which, in turn, bolsters awareness and support for measures needed during health emergency events(3).

The World Health Organization (WHO) is developing guidance on *Learning in Emergencies* to provide evidence-based interventions, training methods and tools for professionals, communities and institutions to ensure quality learning provision and knowledge dissemination during health emergencies. The primary reason to develop the guidance was due to lessons learned from the COVID-19 pandemic The COVID-19 pandemic presented unique circumstances and global challenges when it became clear that ‘just in time’ learning was required to retrain and upskill large numbers of health professionals in order to launch an effective response. For example, new methods and approaches for testing and surveillance at scale were proven essential to operate at the national, regional, and local level, which involved the need for rapid dissemination and uptake of new learning so that the entire health, social care and education workforce were equipped with relevant knowledge and skills to respond effectively, especially in low and middle income countries(4, 5).

The COVID-19 pandemic also prompted the need to integrate effective risk communication strategies to tackle the evolving information needs and misinformation or infodemic (i.e., short for information epidemic) (6) and potential spread of misinformation that could negatively impact on efforts to develop and deliver learning interventions to professionals, communities and institutions(7). From the onset of the COVID-19 pandemic, WHO committed to knowledge dissemination to frontline health workers, governmental and non-governmental actors, policy makers, capacity builders and trainers as well as the public via its low-bandwidth adjusted online platform. In addition to regular updates as more research emerged, WHO intended to regularly assess the effectiveness of this initiative - that is knowledge dissemination through its online platform (OpenWHO) on all the aspects of tackling the pandemic and then including other health emergencies. Key findings indicated that employing strategies to make learning more equitable and accessible can yield better results in terms of outreach, and that learning production must be targeted for real-time events in languages spoken in outbreak impacted areas(8). Nonetheless, learning design based on inclusive pedagogy and learning sciences, while being cognizant of barriers, such as poor internet connection or limited digital literacy, to access learning can optimize learning experience(9).

The second rationale for developing the Learning in Emergencies guidance is that various current guidelines tackled adult learning partially or in one dimension. WHO itself has published several frameworks and recommendations. However, none of these related guidelines covers Adult Learning in Health Emergencies. The Learning in Emergencies guidance will address a priority issue cited during the World Health Assembly (WHA), to help WHO support governments in their health-related capacity building and to reach health learning goals. The *Learning in Emergencies* guidance will encompass the full scope of learning as preparedness, readiness, response and resilience actions in capacitation for public health emergencies.

This scoping review of literature was undertaken to inform further methodological choices and the commissioning of subsequent systematic reviews to feed into the Guidance development process.

### Objectives

The objective of this scoping review was to map the existing evidence (including qualitative, quantitative and mixed-methods peer-reviewed publications, grey literature studies, systematic and other types of literature reviews and qualitative evidence synthesis) that has been published on the topic.

### Target population

The target population included experts and individuals in need of health information, such as health workforce, experts and volunteers, national institutions and ministries (governmental and non-governmental actors), policymakers, academia, capacity builders and trainers, citizens and affected populations.

### Phenomenon of interest

The phenomena of interest were classified as learning interventions as specified below and health emergencies including the follow types: Biological, Environmental, Armed conflicts, Deliberate acts of terrorism and Industrial accidents.

### Intervention/Exposure

Learning interventions included:

- Continuous learning for professionals preparing for or acting in health emergencies
- Adult learning interventions and methods in emergency situations
- Professional education, training and learning
- Real time, just in time learning
- Knowledge and learning transfer from an expert organization
- Learning readiness in anticipation and preparedness for any health hazard

## Methods

The methodology was guided by Arksey and O’Malley’s (8) five stage framework for scoping reviews: identifying the research question(s), identifying relevant studies, charting the data, collating, summarizing, and reporting the results. In addition, principles of mixed-methods framework synthesis were used to manage diverse study designs and help extract, map, chart, categorize and summarize study findings(10). The review was reported using the relevant domains of the Preferred Reporting items for Systematic Review and Meta-Analysis for scoping reviews (PRISMA-ScR)(11).

A priori protocol was developed and published on the Open Science Framework: https://doi.org/10.17605/OSF.IO/5BK9R.

### Selecting relevant studies

Searches were carried out between February 2nd and February 28th 2023, and covered sources from 2003 to the present. Key search terms were identified within three PICO question components (Table 1 Supplementary material). These terms were also used to identify relevant documents from which Medical Subject Heading (MeSH) or other database-specific terms and keywords could be extracted.

Key terms were searched using a free text strategy in the titles and abstracts. This allowed having a broader, more sensitive approach and eliminated the possibility of relevant items being missed. MeSH was applied to give more specific results.

The following databases were searched: Pubmed/Medline and Embase, using predefined combinations of key search terms. To prioritize LMICs, filters of the selected databases were applied. The search strategy and search words are annexed (Table 2 Supplementary material).

Search results were scanned for relevance and those meriting further examination were imported into Rayyan for further consideration. The search and initial screening were undertaken by GB and studies checked by other authors were included. Citations were excluded if they focused exclusively on academic settings (early education through medical school), did not have a learning intervention (for example studies addressing knowledge in unprepared professionals) or were not contextualized during or for a health emergency. News (or announcements) as well as protocols of studies/reviews that had not been completed, and other irrelevant document types were excluded, as well as non-peer reviewed articles.

Titles and abstracts were screened against the following inclusion criteria: (1) published between 2003 and 2023; (2) abstract and title written in English; and (3) presenting data on research question (date, methodology, with focus on geographical low and middle income context, type of learning/intervention and for whom, type of emergency), (4) peer-reviewed sources only. Then, screening of full texts of pre-selected citations against above-mentioned criteria was performed. At this stage full texts written in other UN-languages and Portuguese language were included as well. Processes were undertaken by GB and crossed checked by co-authors. We did not undertake double blind processing.

### Charting the data

All the included evidence was extracted into an Excel spreadsheet and included citations were exported into Endnote. Data were extracted systematically using a standardized form that included information on the period of study, location, study population, design, research questions, key findings, and conclusions.

At the mapping stage we did not focus on the findings and conclusions as not all studies were selected for inclusion in the systematic reviews that will subsequently be commissioned.

### Collating, summarizing and reporting results

The charted data (in the form of an evidence map) were then further analyzed, grouped and sorted guided by the review aim and objectives. We specifically focused on developing an understanding of the available literature on the phenomena of interest and created visual displays and tables. We also identified and described four major topic areas.

## Results

A total of 6411 articles were imported into Rayyan, of which 1656 were duplicates and 4317 were excluded based on a brief scan of titles and abstracts. The remaining 446 documents were sorted by full-text access. One hundred and thirteen articles were excluded for not being relevant and fourteen did not have full text access. As shown in Fig 1, 319 articles met the inclusion criteria and were included. Full description of included studies can be found in Table 3 of the Supplementary material.

**Fig 1:**
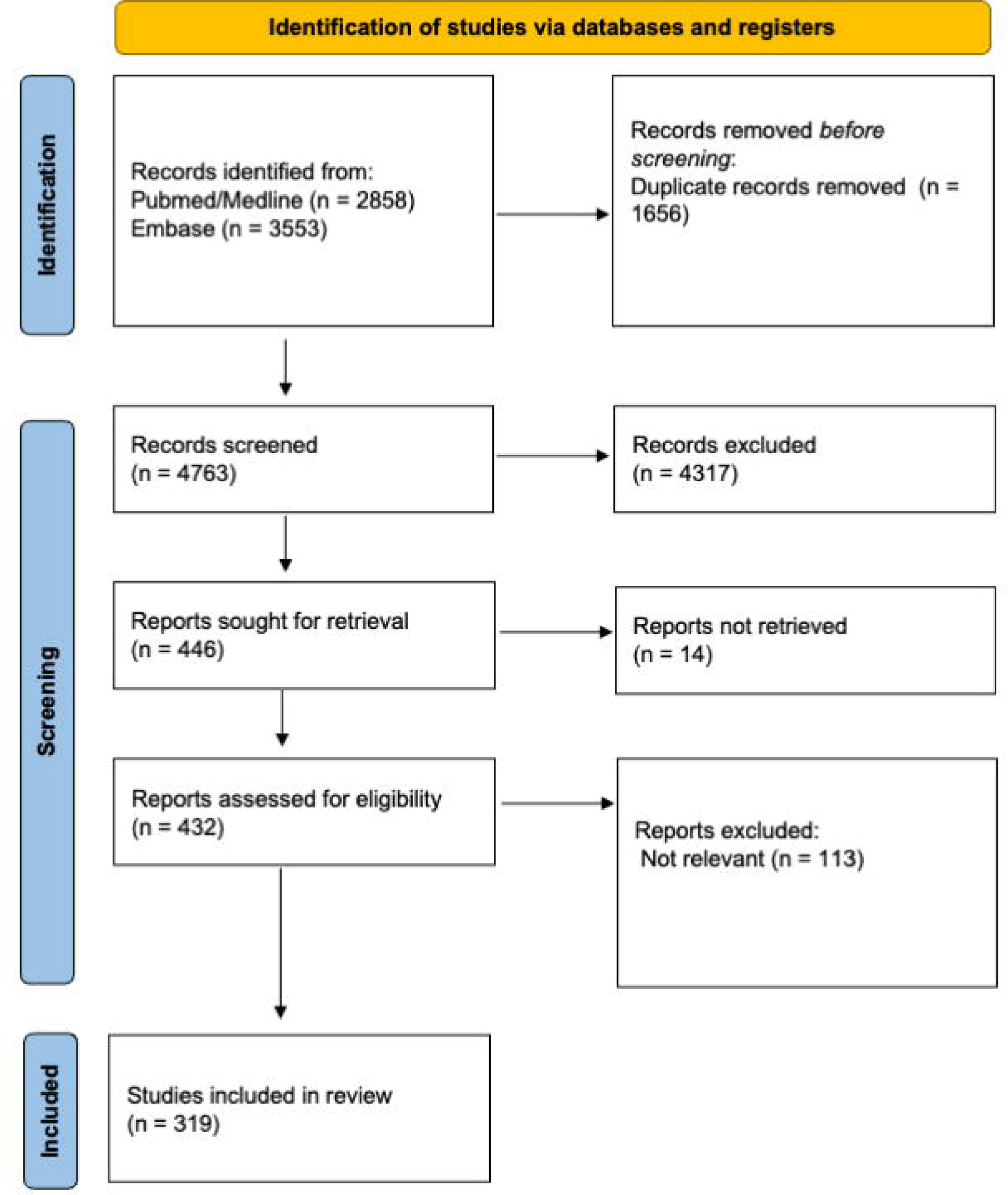
PRISMA Flowchart

### Overview of study characteristics

Most of the articles were descriptive studies on experiences in different countries(12–176), followed by before-after studies(169, 173, 177–260). Also, we found 19 randomized controlled trials(261–279), 14 cross-sectional studies(280–293), 12 observational studies(294–305), 7 reviews(306–312), 5 qualitative studies(313–317) and 3 opinion pieces(318–320). (Table 4 Supplementary material)

### Emergency context

COVID-19, Disasters in general, Ebola and wars were the most frequent topics. This is related to the publication dates because most studies have been published during these health emergencies (Ebola, 2015 and COVID-19 during 2020, 2021 and 2022) (Fig 2)

**Fig 2:**
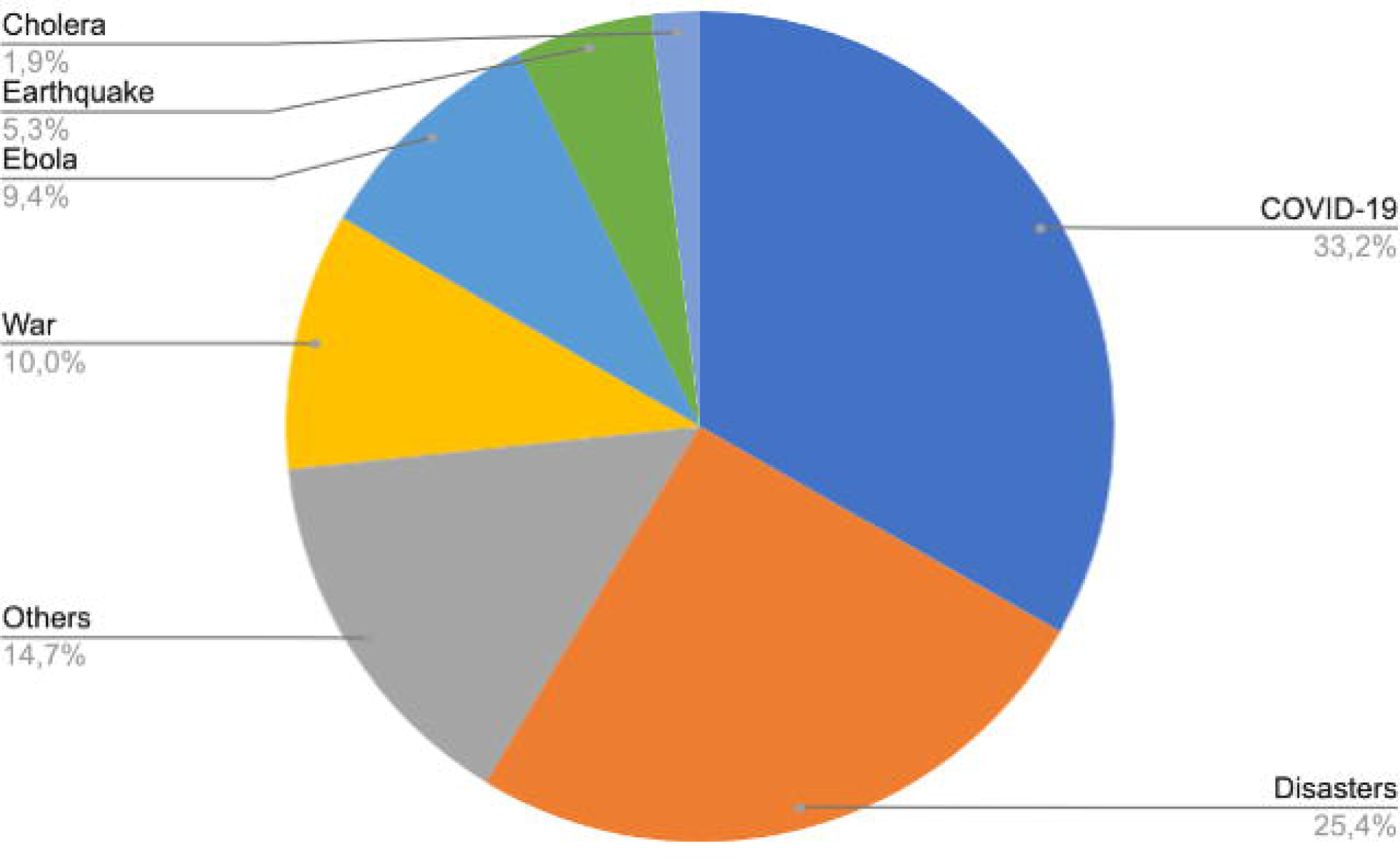
Type of emergency distribution

### Location

Most studies were conducted in China (mainly due to COVID-19 and Earthquakes), India and South Africa. A large number of the learning evidence was also derived from several countries in Africa.

### Interventions, learning methods and tools

Different types of learning interventions are described across the studies. The majority of the studies used in-person training as an intervention(12, 14, 18–22, 30, 33, 34, 37–42, 46, 48–50, 52, 55, 61, 66, 67, 70, 72, 73, 75–78, 80–82, 87, 93, 97–100, 104, 108, 109, 111, 118, 120, 122–126, 128–131, 133, 135, 136, 138–141, 143, 144, 146–151, 153–159, 161, 165, 172, 174, 178–182, 184, 185, 187–189, 193, 195, 197, 198, 200, 206, 210, 212, 214–217, 220, 222, 223, 225, 228, 229, 232, 234, 237, 238, 243–245, 247–254, 257, 259, 261, 262, 264, 266, 268, 269, 271, 273, 275, 277, 279, 282, 288–290, 292, 294, 295, 298, 302, 313, 315, 317, 318, 320, 321), followed by virtual training.

Lectures, discussions, role playing, ‘hands on’ basic skills training, materials, videos and simulations were used as in-person modalities. Additionally, 9 studies(26, 83, 85, 89, 117, 224, 263, 297, 307) suggested a professional approach (such as Masters degrees and post graduate studies in universities) as a way of preparing healthcare workers for health emergencies.

Within the virtual modalities, studies describe the use of Telemedicine(91, 92, 94, 96, 192, 283, 289), Massive Online Open Courses (MOOC)(28, 166, 208, 280, 296), Social Media(15, 265, 281, 284), Gamification(183, 274), Virtual simulation(88, 272), Artificial Intelligence(86, 300) and Mobile devices(147, 202).

A few studies used blended format, usually workshops followed by virtual training (in-person and virtual)(35, 44, 54, 63, 64, 101, 126, 127, 152, 191, 241, 270, 301, 318–320) (Fig 3)

**Fig 3:**
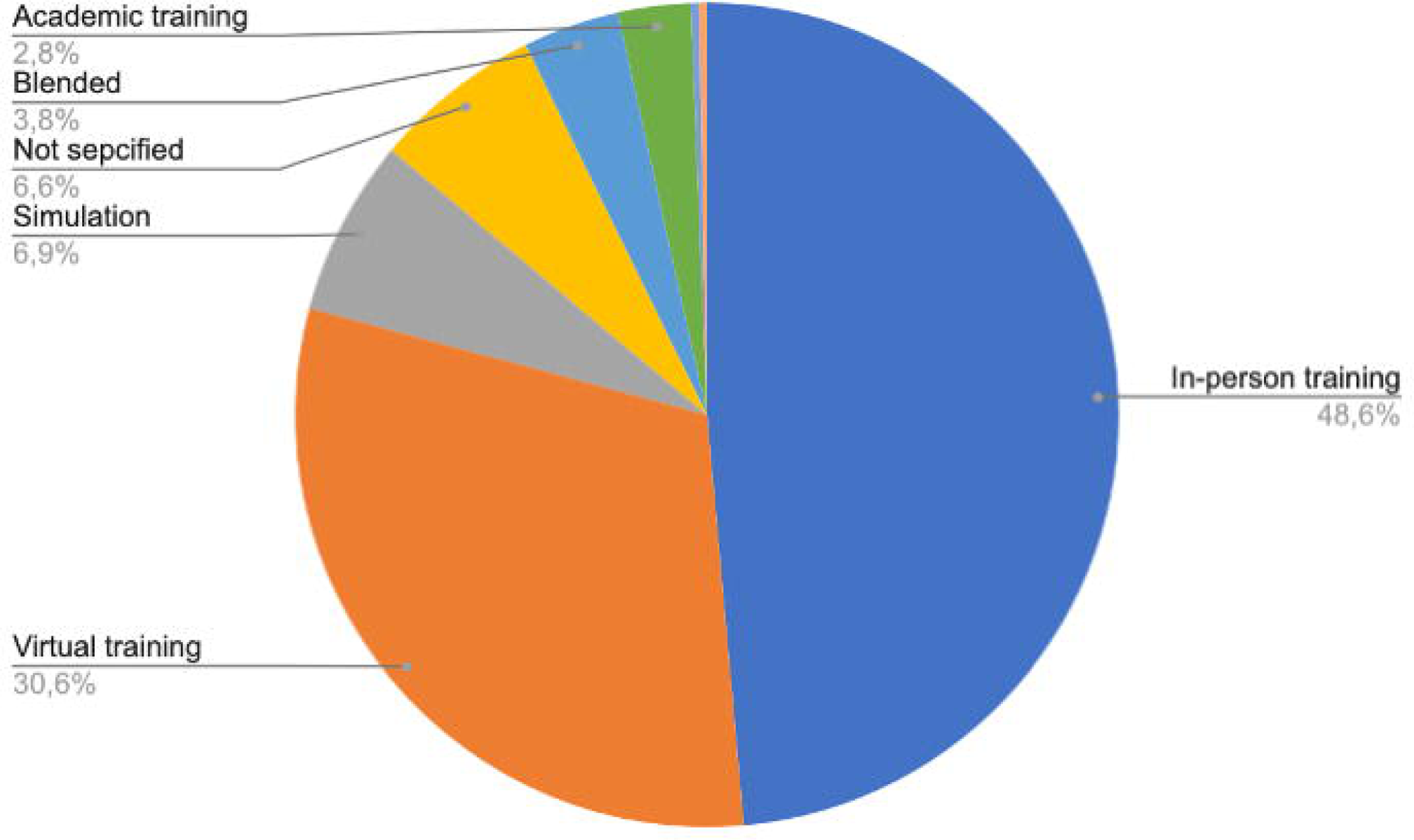
Type of learning method distribution

### Learners

Most of the learning interventions were directed at the health workforce such as medical doctors, nurses, dentists, medical students, laboratory staff and paramedical students. Some learning interventions were also aimed at military personnel, citizens and affected population, volunteers, academia and national institutions. (Table 5 Supplementary material)

### Learning content

Curricula content was mostly focused on disease and disaster management. Some content was about personal safety and security(48, 49, 54, 203–205, 207, 208), waste management(65, 211, 298), mental health(94, 104, 165, 171, 176, 212, 220, 234, 258, 276, 292), infection control and prevention(138, 167, 240, 263, 301), pest control(64, 65, 69, 211, 215, 298), triage(15, 61, 90, 93, 206, 270, 289) and stress management(30, 181, 269). A smaller number of curricula included surgical training(97, 98, 164, 322), time management(222), stigma(268), language and local culture(19), humanitarian law and leadership(101), chemical incidents(213) and dealing with ethical dilemmas(71).

### Major topic areas

By grouping the studies, four major topics were identified:

1. Knowledge acquisition: Articles that analyzed how much knowledge was acquired during learning interventions as a way to assess effectiveness of the intervention methods.
2. Emergency Plans: Learning recommendations focusing on how governments and institutions should be prepared to face health emergencies.
3. Impact of the learning intervention: Articles that analyzed how learning and training during health emergencies had an impact on trainees and affected populations.
4. Training method: Articles that focus on different learning methods and tools that can be used for knowledge transfer during a health emergency.

The distribution of the major topics across the evidence is shown in Fig 4.

**Fig 4:**
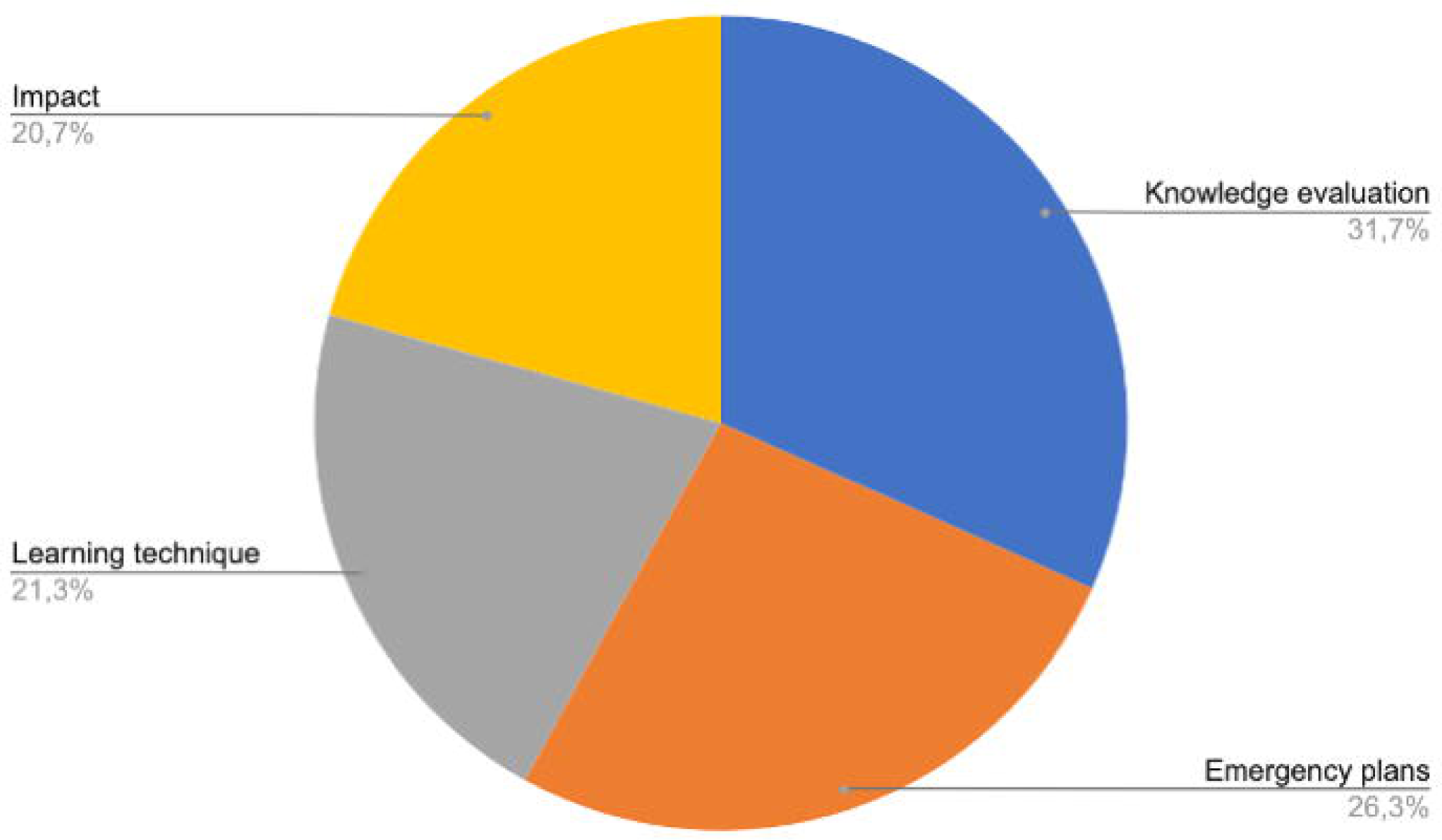
Major topics distribution

The largest number of articles found (n = 101) correspond to “Knowledge evaluation”, followed by articles corresponding to “Emergency Plans” (n=84) and “Training Method” (n=68). Finally, “Impact” was analyzed in 66 studies.

### Knowledge acquisition

For knowledge evaluation, 101 studies(17, 18, 33, 41, 42, 48, 65, 79, 90, 94, 95, 97, 99, 100, 103, 107, 109, 111, 116, 155, 163, 168, 169, 172, 175, 177–180, 182, 185–187, 189, 194–198, 200, 202, 205–207, 211, 212, 214, 216, 217, 219, 221, 223–228, 230–235, 237, 239, 243–245, 248, 249, 252, 253, 256, 258, 259, 264–266, 268, 270, 272, 273, 278, 279, 283, 284, 286, 288–290, 292, 294, 298, 300–302, 304, 305, 315, 317) provided information about how a learning intervention during a health emergency can impact the knowledge of trainees. According to the results of these studies, learning interventions were helpful for improving knowledge, preparedness and confidence of trainees. On the other hand, improved knowledge had a positive impact on diagnosis, and health outcomes such as rate of infected patients.

### Emergency Plans

84 studies(25, 35, 37–39, 43, 49–51, 53, 54, 56, 59, 60, 62, 63, 66, 68, 70, 72–74, 77, 80–82, 85, 102, 106, 112, 113, 117, 118, 120, 121, 125–128, 130–134, 136, 141, 146, 148, 149, 151, 154, 156–158, 161, 162, 164, 165, 171, 173, 174, 176, 183, 190, 238, 247, 255, 262, 267, 274, 285, 287, 306–311, 316, 318, 319, 323–325) described the use of learning interventions as part of their emergency plans from countries and health institutions: 65 are reports, 6 are reviews and the rest of the evidence are randomized control trials, before and after studies, opinions, cross-sectional and qualitative studies. The evidence described how countries and hospitals should train healthcare workers, military, citizens and students in environmental emergencies (earthquake, tsunami, typhoons and disasters in general), biological emergencies (COVID-19, HIV, syphilis, Ebola, Cholera and Zika) and armed conflicts (war and chemical emergencies). The objective of this training was to prepare different populations for a future emergency and this explains why these interventions were not offered during an emergency.

As for the RCT, Ma et al(274) compared the use of a gaming technique versus a disaster simulation and Lee et al(267) compared two video techniques: basic response to a fire versus generic volcanic emergency. In both studies, knowledge competence and response were statistically higher in the intervention groups (gaming technique and video response to fire emergencies).

### Training Methods

On training methods, 68 studies(12–16, 19–24, 26–29, 31, 32, 36, 40, 44, 46, 47, 52, 57, 58, 61, 64, 67, 78, 83, 84, 86, 87, 89, 92, 96, 110, 122–124, 137, 140, 144, 145, 147, 152, 160, 166, 167, 169, 192, 203, 213, 215, 246, 250, 254, 260, 293, 296, 297, 299, 303, 312, 320, 321, 326) focused on describing different training methods that can be used during a health emergency. 15 studies described in-person training methods, and the rest of them used a virtual training method such as simulation, telemedicine, MOOC, videos or artificial intelligence.

MOOCs were used for cholera(28) and COVID-19(166, 296, 314), and all of these studies showed that this method can be useful to disseminate trustworthy information in low and middle-income countries and fill the gap of information during health emergencies.

Artificial intelligence(86) was used during COVID-19 to help radiologists to efficiently and timely diagnose the COVID-19 suspected patients.

Videos(203) were implemented in Burkina Faso during a dengue epidemic. This study concluded that videos are effective for knowledge transfer and training health professionals, however the narrative genre of the videos can influence knowledge acquisition.

### Impact of learning interventions

Two studies measured impact as satisfaction in citizens and affected populations: One of them (261) trained parents of high-risk children about malaria prevention using an in-person technique. Participant satisfaction was assessed using a qualitative approach, however the main challenge reported was adherence to the course. The other study(184) reported on in-person training of Syrian refugee mothers of children with autism about overcoming trauma by war. Satisfaction was analyzed and improvements were recommended by the attendees, such as using online methods during the course.

In the rest of the evidence that reported satisfaction as an outcome, it was measured as self-perceived, as mental health status of healthcare workers, well-being and preparedness.

27 studies analyzed how training during a health emergency impacted on the quality of life of healthcare workers, citizens and volunteers. The endpoints included: acceptability(295), anxiety(209), confidence(193), mental health (45, 199, 220, 222, 276, 313), preparedness(135, 139, 142, 143, 153, 159, 257, 269, 277, 327), satisfaction(30, 170, 198, 328–330), community cohesion(269) and social adaptation(291). Most of the learning interventions were given after health emergencies occurred. This would be related to the fact that quality of life of the workers would be affected after the occurrence of these events. Two studies(193, 209), however, analyzed how quality of life could be improved by receiving prior training.

## Discussion

We found 319 studies that analyzed different learning interventions and training methods during health emergencies. Information about virtual training (including online platforms, artificial intelligence, MOOC, the use of mobile devices, social networks and telemedicine) arose mainly during the COVID-19 pandemic. Before the pandemic, most of the studies focused on in-person training. This change could be explained by technological advancements, the need for new information that had to be updated quickly and also because of social distancing recommendations. Of note, access to new technologies was scaled up after 2020 as the pandemic accelerated this process.

Many studies analyzed the impact of training on clinical outcomes such as patient survival (19, 215), rate of workers infection(33, 41, 87), number of hospitalizations(99, 299) and number of correct diagnoses(86, 300). These findings will be important when developing guidance since these outcomes can be useful for decision making and selection of appropriate learning interventions and methods. Finally, the importance of coordinated work between different institutions(16, 25, 28, 38, 112, 238, 285), universities, governments and non-governmental organizations emerges was noted in several studies. How best to foster coordinated work between institutions may be of great interest for future research.

We also found some potential evidence gaps that are worth investigating in future research studies. First, there is limited evidence on the content of learning interventions, especially regarding ethical dilemmas and how to solve them (71). Also, we found a scarcity of evidence on end-of-life management during emergencies such as end-of-life care and managing deceased people. Subsequently, we identified other evidence gaps such as lack of robust evaluation of the effect of learning transfer, as most studies rely on self-reports (also referred as post-training ‘smile-sheets’) to evaluate transfer of learning(331–333). These studies provide limited actionable data to determine the effectiveness of training programs, as getting a favorable reaction from learners does not guarantee that learning transfer has been achieved(334). Further, too little research has examined the accountability of trainers for transfer in terms of using transfer-enhancing strategies, and how trainers are being evaluated(335) In addition, most studies focused on information dissemination to build skills rather than employing evidence-informed training methods for performance improvement at the time of emergencies. Poor learning design and evaluation methods can result in learning being wasted(331). Lastly, there was a lack of evidence on managing emergencies through teamwork, effective communication, and stress management. After all, factors such as stress, poor team dynamic due to hierarchical issues, and poor communication can lead to morbidity or mortality.

## Limitations and strengths

This scoping review has potential limitations and strengths. One such limitation was the time constraint to deliver the scoping review quickly. Despite following standard scoping review processes, the search was not as robust as when performing a systematic review. However, once the *Learning in Emergencies* guidance is under way and new systematic reviews on the subject are commissioned, new evidence may arise to complement these findings. Scoping reviews are limited in purpose, but often fulfill a vital first step in establishing the amount and type of evidence from which subsequent systematic reviews can be commissioned with greater certainty of feasibility. The scoping review followed an a priori protocol and was undertaken by a diverse team with varied expertise in the topic and scoping review methods.

## Conclusion

Research on learning and learning dissemination during health emergencies has revealed considerable advancements, particularly in virtual learning. Overall, it is evident that learning during health emergencies appears to improve knowledge, management, quality of life, satisfaction and clinical outcomes. All the information provided by this review can give decision makers tools to select different types of learning interventions and methods for healthcare workers, volunteers, military, civilians and governments. This scoping review may also be useful for future research to address the identified evidence gaps.

## Supporting information

PRISMA

## Data Availability

All relevant data are within the manuscript and its Supporting Information files.

## Acknowledgments

The authors thank Aphaluck Bhatiasevi for reviewing this paper and the support of the Learning and Capacity Development Unit, the WHO Health Emergencies Programme and the World Health Organization.

## Funding

This study did not receive any funding.

## Notes

### Competing Interest Statement

The authors have declared no competing interest.

### Clinical Protocols

https://doi.org/10.17605/OSF.IO/5BK9R

### Summary of Updates

Funding information

