## Supplementary Material for "Learning interventions and training methods in health emergencies: A scoping review"

Table 1. PICO question

| **Population** | **Intervention** | **Phenomenon** |
| --- | --- | --- |
| **(Health Personnel[Mesh]** OR Health Personnel[tiab] OR Healthcare Personnel[tiab] OR Health Worker*[tiab] OR Nurse*[tiab] OR Doctor*[tiab] OR Physician*[tiab] OR Paramedic*[tiab] OR Caregiver*[tiab] OR Care Giver*[tiab] OR Medic*[tiab) | **(Education, Distance[Mesh]** OR Distance Education[tiab] OR Online Education[tiab] OR ELearning[tiab] OR Learning[tiab] OR Workshop*[tiab] OR Training[tiab]) | **(Disasters[Mesh]** OR Disaster*[tiab] OR Catastroph*[tiab] OR Mass Casualt*[tiab] OR Terroris*[tiab] OR Bioterroris*[tiab] OR **Epidemics[Mesh**] OR Epidemic*[tiab] OR Pandemic*[tiab] OR Natural Hazard*[tiab] OR Humanitarian Emergen*[tiab] OR Avalanche*[tiab] OR Storm*[tiab] OR Cyclon*[tiab] OR Drought*[tiab] OR Hurricane*[tiab] OR Typhoon*[tiab] OR Earthquake*[tiab] OR Volcanic*[tiab] OR **Volcanic Eruptions**[Mesh] OR Volcanic*[tiab] OR Volcano*[tiab] OR Tsunami*[tiab] OR Flood*[tiab] OR Wildfire*[tiab] OR Wild-Fire*[tiab] OR Wildfire*[tiab] OR Bushfire*[tiab] OR Landslide*[tiab] OR Industrial Accident*[tiab] OR Chemical Hazard*[tiab] OR Chemical Emergenc*[tiab] OR Nuclear Hazard*[tiab] OR Nuclear Emegenc*[tiab] OR Radiological Hazard*[tiab] OR Radiological Emergenc*[tiab] OR Biological Hazard*[tiab] OR Biological Emergenc*[tiab] OR Biohazard*[tiab] OR Warfare*[tiab] OR Armed Conflict*[tiab] OR War[tiab]) |

Table 2. Search strategy

Pubmed search

| **Search** | **Actions** | **Details** | **Query** | **Results** | **Time** |
| --- | --- | --- | --- | --- | --- |
| #4 |  |  | **#1 AND #2 AND #3** | [14,186](https://pubmed.ncbi.nlm.nih.gov/?term=%231+AND+%232+AND+%233&sort=) | 10:41:57 |
| #3 |  |  | **(Disasters[Mesh] OR Disaster*[tiab] OR Catastroph*[tiab] OR Mass Casualt*[tiab] OR Terroris*[tiab] OR Bioterroris*[tiab] OR Epidemics[Mesh] OR Epidemic*[tiab] OR Pandemic*[tiab] OR Natural Hazard*[tiab] OR Humanitarian Emergen*[tiab] OR Avalanche*[tiab] OR Storm*[tiab] OR Cyclon*[tiab] OR Drought*[tiab] OR Hurricane*[tiab] OR Typhoon*[tiab] OR Earthquake*[tiab] OR Volcanic*[tiab] OR Volcanic Eruptions[Mesh] OR Volcanic*[tiab] OR Volcano*[tiab] OR Tsunami*[tiab] OR Flood*[tiab] OR Wildfire*[tiab] OR Wild-Fire*[tiab] OR Wildfire*[tiab] OR Bushfire*[tiab] OR Landslide*[tiab] OR Industrial Accident*[tiab] OR Chemical Hazard*[tiab] OR Chemical Emergenc*[tiab] OR Nuclear Hazard*[tiab] OR Nuclear Emegenc*[tiab] OR Radiological Hazard*[tiab] OR Radiological Emergenc*[tiab] OR Biological Hazard*[tiab] OR Biological Emergenc*[tiab] OR Biohazard*[tiab] OR Warfare*[tiab] OR Armed Conflict*[tiab] OR War[tiab])** | 603,360 | 10:41:20 |
| #2 |  |  | Search: **(Education, Distance[Mesh] OR Distance Education[tiab] OR Online Education[tiab] OR ELearning[tiab] OR Learning[tiab] OR Workshop*[tiab] OR Training[tiab])** | [936,830](https://pubmed.ncbi.nlm.nih.gov/?term=%28Education%2C+Distance%5BMesh%5D+OR+Distance+Education%5Btiab%5D+OR+Online+Education%5Btiab%5D+OR+ELearning%5Btiab%5D+OR+Learning%5Btiab%5D+OR+Workshop%2A%5Btiab%5D+OR+Training%5Btiab%5D%29&sort=) | 10:39:18 |
| #1 |  |  | Search: **(Health Personnel[Mesh] OR Health Personnel[tiab] OR Healthcare Personnel[tiab] OR Health Worker*[tiab] OR Nurse*[tiab] OR Doctor*[tiab] OR Physician*[tiab] OR Paramedic*[tiab] OR Caregiver*[tiab] OR Care Giver*[tiab] OR Medic*[tiab)** | [3,385,086](https://pubmed.ncbi.nlm.nih.gov/?term=%28Health+Personnel%5BMesh%5D+OR+Health+Personnel%5Btiab%5D+OR+Healthcare+Personnel%5Btiab%5D+OR+Health+Worker%2A%5Btiab%5D+OR+Nurse%2A%5Btiab%5D+OR+Doctor%2A%5Btiab%5D+OR+Physician%2A%5Btiab%5D+OR+Paramedic%2A%5Btiab%5D+OR+Caregiver%2A%5Btiab%5D+OR+Care+Giver%2A%5Btiab%5D+OR+Medic%2A%5Btiab%29&sort=) \| 10:38:55 |  |

The filter for Low and Middle-income countries was then applied. The final search strategy is presented below:

**Full Strategy:**

**(Disasters[Mesh]** OR Disaster*[tiab] OR Catastroph*[tiab] OR Mass Casualt*[tiab] OR Terroris*[tiab] OR Bioterroris*[tiab] OR **Epidemics[Mesh**] OR Epidemic*[tiab] OR Pandemic*[tiab] OR Natural Hazard*[tiab] OR Humanitarian Emergen*[tiab] OR Avalanche*[tiab] OR Storm*[tiab] OR Cyclon*[tiab] OR Drought*[tiab] OR Hurricane*[tiab] OR Typhoon*[tiab] OR Earthquake*[tiab] OR Volcanic*[tiab] OR **Volcanic Eruptions**[Mesh] OR Volcanic*[tiab] OR Volcano*[tiab] OR Tsunami*[tiab] OR Flood*[tiab] OR Wildfire*[tiab] OR Wild-Fire*[tiab] OR Wildfire*[tiab] OR Bushfire*[tiab] OR Landslide*[tiab] OR Industrial Accident*[tiab] OR Chemical Hazard*[tiab] OR Chemical Emergenc*[tiab] OR Nuclear Hazard*[tiab] OR Nuclear Emegenc*[tiab] OR Radiological Hazard*[tiab] OR Radiological Emergenc*[tiab] OR Biological Hazard*[tiab] OR Biological Emergenc*[tiab] OR Biohazard*[tiab] OR Warfare*[tiab] OR Armed Conflict*[tiab] OR War[tiab]) **AND** **(Health Personnel[Mesh]** OR Health Personnel[tiab] OR Healthcare Personnel[tiab] OR Health Worker*[tiab] OR Nurse*[tiab] OR Doctor*[tiab] OR Physician*[tiab] OR Paramedic*[tiab] OR Caregiver*[tiab] OR Care Giver*[tiab] OR Medic*[tiab) **AND** **(Education, Distance[Mesh]** OR Distance Education[tiab] OR Online Education[tiab] OR ELearning[tiab] OR Learning[tiab] OR Workshop*[tiab] OR Training[tiab]) **AND** (LMIC[tiab] OR "Low and Middle"[tiab] OR Subsaharian[tiab] OR Sub Saharian[tiab] OR Southeast Asia*[tiab] OR Middle East*[tiab] OR Central America*[tiab] OR Africa[Mesh] OR Afghanistan[Mesh] OR Afghan*[tiab] OR Benin[Mesh]OR Benin*[tiab] OR Burkina Faso[Mesh] OR Burkin*[tiab] OR Burundi[Mesh] OR Burundi*[tiab] OR Central African Republic[Mesh] OR Central African[tiab] OR Chad[Mesh] OR Chad[tiab] OR Albania[Mesh] OR Albania*[tiab] OR Angola[Mesh] OR Angola[tiab] OR Algeria[Mesh] OR Algeria*[tiab] OR Armenia[Mesh] OR Armenia*[tiab] OR Azerbaijan[Mesh] OR Azerbaijan*[tiab] OR Bangladesh[Mesh] OR Bangladesh*[tiab] OR Republic of Belarus[Mesh] OR Belarus*[tiab] OR Belize[Mesh] OR Beliz*[tiab] OR Bhutan[Mesh] OR Bhutan*[tiab] OR Bolivia[Mesh] OR Bolivia*[tiab] OR “Bosnia and Herzegovina”[Mesh] OR Bosni*[tiab] OR Botswana[Mesh] OR Botswan*[tiab] OR Brazil[Mesh] OR Brazil*[tiab] OR Bulgaria[Mesh] OR Bulgaria*[tiab] OR Cabo Verde[Mesh] OR Cabo Verde*[tiab] OR Cambodia[Mesh] OR Cambodia*[tiab] OR Cameroon[Mesh] OR Cameroon*[tiab] OR China[Mesh] OR China[tiab] OR Chinese[tiab] OR Colombia[Mesh] OR Colombia*[tiab] OR Comoros[Mesh] OR Comoro*[tiab] OR Democratic Republic of the Congo[Mesh] OR Congo*[tiab] OR Costa Rica[Mesh] OR Costa Rica[tiab] OR Costarica*[tiab] OR Cote d'Ivoire[Mesh] OR “Côte d'Ivoire”[tiab] OR Cuba[Mesh] OR Cuba*[tiab] OR Djibouti[Mesh] OR Djibout*[tiab] OR Dominican Republic[Mesh] OR Dominic*[tiab] OR Ecuador[Mesh] OR Ecuador*[tiab] OR Egypt[Mesh] OR Egypt*[tiab] OR El Salvador[Mesh] OR Salvador*[tiab] OR Eritrea[Mesh]OR Eritrea*[tiab] OR Ethiopia[Mesh] OR Ethiopi*[tiab] OR Fiji[Mesh] OR Fiji*[tiab] OR Gabon[Mesh] OR Gabon*[tiab] OR Gambia[Mesh] OR Gambia*[tiab] OR “Georgia (Republic)”[Mesh] OR Georgia*[tiab] OR Ghana[Mesh] OR Ghana*[tiab] OR Guatemala[Mesh] OR Guatemal*[tiab] OR Guinea[Mesh] OR Guinea-Bissau[Mesh] OR Guinea*[tiab] OR Guyana[Mesh] OR Guyan*[tiab] OR Gabon OR Haiti[Mesh] OR Haiti*[tiab] OR Honduras[Mesh] OR Hondur*[tiab] OR India[Mesh] OR India[tiab] OR Indonesia[Mesh] OR Indones*[tiab] OR Iran[Mesh] OR Iran*[tiab] OR Iraq[Mesh] OR Iraq[tiab] OR Jamaica[Mesh] OR Jamai*[tiab] OR Jordan[Mesh] OR Jordan*[tiab] OR Kazakhstan[Mesh] OR Kazakhstan*[tiab] OR Kenya[Mesh] OR Kenya*[tiab] OR Micronesia[Mesh] OR Micronesia*[tiab] OR Kiribati*[tiab] OR Kosovo[Mesh] OR Kosov*[tiab] OR Kyrgyzstan[Mesh] OR Kyrgyzstan*[tiab] OR “Democratic People's Republic of Korea”[Mesh] OR North Korea*[tiab] OR Laos[Mesh] OR Laos*[tiab] OR Lebanon[Mesh] OR Leban*[tiab] OR Lesotho[Mesh] OR Lesoth*[tiab] OR Liberia[Mesh] OR Liberia*[tiab] OR Libya[Mesh] OR Libya*[tiab] OR “Macedonia (Republic)”[Mesh] OR Macedonia*[tiab] OR Madagascar[Mesh] OR Madagascar*[tiab] OR Malawi[Mesh] OR Malawi*[tiab] OR Mali[Mesh] OR Mali[tiab] OR Mauritania[Mesh] OR Mauritan*[tiab] OR Mauritius[Mesh] OR Mauriti*[tiab] OR Mexico[Mesh] OR Mexic*[tiab] OR Moldova[Mesh] OR Moldov*[tiab] OR Mongolia[Mesh] OR Mongolia*[tiab] OR Montenegro[Mesh] OR Montenegr*[tiab] OR Morocco[Mesh] OR Morocc*[tiab] OR Myanmar[Mesh] OR Myanmar*[tiab] OR Mozambique[Mesh] OR Mozambiq*[tiab] OR Namibia[Mesh] OR Namibia*[tiab] OR Nepal[Mesh] OR Nepal*[tiab] OR Nicaragua[Mesh] OR Nicaragu*[tiab] OR Niger[Mesh] OR Niger*[tiab] OR Nigeria[Mesh] OR Nigeri*[tiab] OR Pakistan[Mesh] OR Pakistan*[tiab] OR Palau[Mesh] OR Palau*[tiab] OR Panama[Mesh] OR Panam*[tiab] OR Papua New Guinea[Mesh] OR Papua*[tiab] OR Paraguay[Mesh] OR Paraguay*[tiab] OR Peru[Mesh] OR Peru*[tiab] OR Philippines[Mesh] OR Philippine*[tiab] OR Rwanda[Mesh] OR Rwand*[tiab] OR Samoa[Mesh] OR Samoa*[tiab] OR Senegal[Mesh] OR Senegal*[tiab] OR Serbia[Mesh] OR Serbia*[tiab] OR Sierra Leone[Mesh] OR Sierra Leon*[tiab] OR Somalia[Mesh] OR Somalia*[tiab] OR South Africa[Mesh] OR South Africa[tiab] OR Southafrica*[tiab] OR Sri Lanka[Mesh] OR Sri Lanka*[tiab] OR Swaziland[Mesh] OR Swaziland*[tiab] OR Sudan[Mesh] OR Sudan*[tiab] OR Syria[Mesh] OR Syria*[tiab] OR Tajikistan[Mesh] OR Tajikistan*[tiab] OR Tanzania[Mesh] OR Tanzania*[tiab] OR Thailand[Mesh] OR Thailand*[tiab] OR Timor-Leste[Mesh] OR Timor*[tiab] OR Tonga[Mesh] OR Tonga*[tiab] OR Togo[Mesh] OR Togo*[tiab] OR Tunisia[Mesh] OR Tunisia*[tiab] OR Turkey[Mesh] OR Turk*[tiab] OR Turkmenistan[Mesh] OR Ukraine[Mesh] OR Ukrain*[tiab] OR Uganda[Mesh] OR Ugand*[tiab] OR Uzbekistan[Mesh] OR Uzbekistan*[tiab] OR Venezuela[Mesh] OR Venezuel*[tiab] OR Vietnam[Mesh] OR Vietnam*[tiab] OR Yemen[Mesh] OR Yemen*[tiab] OR Zambia[Mesh] OR Zambia*[tiab] OR Zimbabwe[Mesh] OR Zimbabwe*[tiab])

Embase search

**(Disasters[Mesh]** OR Disaster*[tiab] OR Catastroph*[tiab] OR Mass Casualt*[tiab] OR Terroris*[tiab] OR Bioterroris*[tiab] OR **Epidemics[Mesh**] OR Epidemic*[tiab] OR Pandemic*[tiab] OR Natural Hazard*[tiab] OR Humanitarian Emergen*[tiab] OR Avalanche*[tiab] OR Storm*[tiab] OR Cyclon*[tiab] OR Drought*[tiab] OR Hurricane*[tiab] OR Typhoon*[tiab] OR Earthquake*[tiab] OR **Volcanic Eruptions**[Mesh] OR Volcanic*[tiab] OR Volcano*[tiab] OR Tsunami*[tiab] OR Flood*[tiab] OR Wildfire*[tiab] OR Wild-Fire*[tiab] OR Bushfire*[tiab] OR Landslide*[tiab] OR Industrial Accident*[tiab] OR Chemical Hazard*[tiab] OR Chemical Emergenc*[tiab] OR Nuclear Hazard*[tiab] OR Nuclear Emergenc*[tiab] OR Radiological Hazard*[tiab] OR Radiological Emergenc*[tiab] OR Biological Hazard*[tiab] OR Biological Emergenc*[tiab] OR Biohazard*[tiab] OR Warfare*[tiab] OR Armed Conflict*[tiab] OR War[tiab]) **AND** **(Health Personnel[Mesh]** OR Health Personnel[tiab] OR Healthcare Personnel[tiab] OR Health Worker*[tiab] OR Nurse*[tiab] OR Doctor*[tiab] OR Physician*[tiab] OR Paramedic*[tiab] OR Caregiver*[tiab] OR Care Giver*[tiab] OR Medic*[tiab) **AND** **(Education, Distance[Mesh]** OR Distance Education[tiab] OR Online Education[tiab] OR ELearning[tiab] OR Learning[tiab] OR Workshop*[tiab] OR Training[tiab]) AND (	)

Embase Classic+Embase <2003 to 2023 February 03>

1	exp disaster/	34462

2	Disaster*.ti,ab.	36012

3	Catastroph*.ti,ab.	42907

4	(Mass adj1 Casualt*).ti,ab.	3249

5	Terroris*.ti,ab.	8344

6	Bioterroris*.ti,ab.	4279

7	exp epidemic/	143913

8	Epidemic*.ti,ab.	161539

9	Pandemic*.ti,ab.	208701

10	(Natural adj1 Hazard*).ti,ab.	805

11	(Humanitarian adj1 Emergen*).ti,ab.	387

12	Avalanche*.ti,ab.	3211

13	Storm*.ti,ab.	29420

14	Cyclon*.ti,ab.	3947

15	Drought*.ti,ab.	26557

16	Hurricane*.ti,ab.	4734

17	Typhoon*.ti,ab.	963

18	Earthquake*.ti,ab.	10619

19	Volcanic*.ti,ab.	5178

20	exp volcano/	4481

21	Volcano*.ti,ab.	3793

22	Tsunami*.ti,ab.	3274

23	Flood*.ti,ab.	24313

24	Wildfire*.ti,ab.	3279

25	Wild-Fire*.ti,ab.	103

26	Bushfire*.ti,ab.	507

27	Landslide*.ti,ab.	1021

28	(Industrial adj1 Accident*).ti,ab.	1735

29	(Chemical adj1 Hazard*).ti,ab.	1766

30	(Chemical adj1 Emergenc*).ti,ab.	117

31	(Nuclear adj1 Hazard*).ti,ab.	45

32	(Nuclear adj1 Emergenc*).ti,ab.	353

33	(Radiological adj1 Hazard*).ti,ab.	538

34	(Radiological adj1 Emergenc*).ti,ab.	385

35	(Biological adj1 Hazard*).ti,ab.	709

36	(Biological adj1 Emergenc*).ti,ab.	46

37	Biohazard*.ti,ab.	1173

38	Warfare*.ti,ab.	6504

39	(Armed adj1 Conflict*).ti,ab.	1769

40	War.ti,ab.	49432

41	or/1-40	671661

42	exp health care personnel/	1983234

43	(Health* adj3 Personnel).ti,ab.	15912

44	(Health* adj3 Worker*).ti,ab.	85974

45	Nurse*.ti,ab.	391356

46	Doctor*.ti,ab.	220614

47	Physician*.ti,ab.	660892

48	Medics.ti,ab.	1620

49	Paramedic*.ti,ab.	14540

50	Caregiver*.ti,ab.	119233

51	(Care adj1 Giver*).ti,ab.	5438

52	or/42-51	2674948

53	exp distance learning/	5901

54	(Distance adj1 Education).ti,ab.	1206

55	(Online adj1 Education).ti,ab.	1980

56	ELearning.ti,ab.	806

57	Learning.ti,ab.	554760

58	Workshop*.ti,ab.	65971

59	Training.ti,ab.	711671

60	or/53-59	1213645

61	(LMIC or "Low and Middle" or Subsaharian or "Sub Saharian" or "Southeast Asia*" or "Middle East*" or "Central America*").tw. or exp Africa/ or exp Afghanistan/ or Afghan*.tw. or exp Benin/ or exp "Burkina Faso"/ or Burkin*.tw. or exp Burundi/ or Burundi*.tw. or exp "Central African Republic"/ or "Central African".tw. or exp Chad/ or Chad.tw. or exp Albania/ or Albania*.tw. or exp Angola/ or Angola.tw. or exp Algeria/ or Algeria*.tw. or exp Armenia/ or Armenia*.tw. or exp Azerbaijan/ or Azerbaijan*.tw. or exp Bangladesh/ or Bangladesh*.tw. or exp "Republic of Belarus"/ or Belarus*.tw. or exp Belize/ or Beliz*.tw. or exp Bhutan/ or Bhutan*.tw. or exp Bolivia/ or Bolivia*.tw. or exp "Bosnia and Herzegovina"/ or Bosni*.tw. or exp Botswana/ or Botswan*.tw. or exp Brazil/ or Brazil*.tw. or exp Bulgaria/ or Bulgaria*.tw. or exp "Cabo Verde"/ or "Cabo Verde*".tw. or exp Cambodia/ or Cambodia*.tw. or exp Cameroon/ or Cameroon*.tw. or exp China/ or China.tw. or Chinese.tw. or exp Colombia/ or Colombia*.tw. or exp Comoros/ or Comoro*.tw. or exp "Democratic Republic of the Congo"/ or Congo*.tw. or exp "Costa Rica"/ or "Costa Rica".tw. or Costarica*.tw. or exp "Cote d'Ivoire"/ or "Côte d'Ivoire".tw. or exp Cuba/ or Cuba*.tw. or exp Djibouti/ or Djibout*.tw. or exp "Dominican Republic"/ or Dominic*.tw. or exp Ecuador/ or Ecuador*.tw. or exp Egypt/ or Egypt*.tw. or exp "El Salvador"/ or Salvador*.tw. or exp Eritrea/ or exp Ethiopia/ or Ethiopi*.tw. or exp Fiji/ or Fiji*.tw. or exp Gabon/ or Gabon*.tw. or exp Gambia/ or Gambia*.tw. or exp "Georgia (Republic)"/ or Georgia*.tw. or exp Ghana/ or Ghana*.tw. or exp Guatemala/ or Guatemal*.tw. or exp Guinea/ or exp Guinea-Bissau/ or Guinea*.tw. or exp Guyana/ or Guyan*.tw. or Gabon.mp. or exp Haiti/ or Haiti*.tw. or exp Honduras/ or Hondur*.tw. or exp India/ or India.tw. or exp Indonesia/ or Indones*.tw. or exp Iran/ or Iran*.tw. or exp Iraq/ or Iraq.tw. or exp Jamaica/ or Jamai*.tw. or exp Jordan/ or Jordan*.tw. or exp Kazakhstan/ or Kazakhstan*.tw. or exp Kenya/ or Kenya*.tw. or exp Micronesia/ or Micronesia*.tw. or Kiribati*.tw. or exp Kosovo/ or Kosov*.tw. or exp Kyrgyzstan/ or Kyrgyzstan*.tw. or exp "Democratic People's Republic of Korea"/ or "North Korea*".tw. or exp Laos/ or Laos*.tw. or exp Lebanon/ or Leban*.tw. or exp Lesotho/ or Lesoth*.tw. or exp Liberia/ or Liberia*.tw. or exp Libya/ or Libya*.tw. or exp "Macedonia (Republic)"/ or Macedonia*.tw. or exp Madagascar/ or Madagascar*.tw. or exp Malawi/ or Malawi*.tw. or exp Mali/ or Mali.tw. or exp Mauritania/ or Mauritan*.tw. or exp Mauritius/ or Mauriti*.tw. or exp Mexico/ or Mexic*.tw. or exp Moldova/ or Moldov*.tw. or exp Mongolia/ or Mongolia*.tw. or exp Montenegro/ or Montenegr*.tw. or exp Morocco/ or Morocc*.tw. or exp Myanmar/ or Myanmar*.tw. or exp Mozambique/ or Mozambiq*.tw. or exp Namibia/ or Namibia*.tw. or exp Nepal/ or Nepal*.tw. or exp Nicaragua/ or Nicaragu*.tw. or exp Niger/ or Niger*.tw. or exp Nigeria/ or Nigeri*.tw. or exp Pakistan/ or Pakistan*.tw. or exp Palau/ or Palau*.tw. or exp Panama/ or Panam*.tw. or exp "Papua New Guinea"/ or Papua*.tw. or exp Paraguay/ or Paraguay*.tw. or exp Peru/ or Peru*.tw. or exp Philippines/ or Philippine*.tw. or exp Rwanda/ or Rwand*.tw. or exp Samoa/ or Samoa*.tw. or exp Senegal/ or Senegal*.tw. or exp Serbia/ or Serbia*.tw. or exp "Sierra Leone"/ or "Sierra Leon*".tw. or exp Somalia/ or Somalia*.tw. or exp "South Africa"/ or "South Africa".tw. or Southafrica*.tw. or exp "Sri Lanka"/ or "Sri Lanka*".tw. or exp Swaziland/ or Swaziland*.tw. or exp Sudan/ or Sudan*.tw. or exp Syria/ or Syria*.tw. or exp Tajikistan/ or Tajikistan*.tw. or exp Tanzania/ or Tanzania*.tw. or exp Thailand/ or Thailand*.tw. or exp Timor-Leste/ or Timor*.tw. or exp Tonga/ or Tonga*.tw. or exp Togo/ or Togo*.tw. or exp Tunisia/ or Tunisia*.tw. or exp Turkey/ or Turk*.tw. or exp Turkmenistan/ or exp Ukraine/ or Ukrain*.tw. or exp Uganda/ or Ugand*.tw. or exp Uzbekistan/ or Uzbekistan*.tw. or exp Venezuela/ or Venezuel*.tw. or exp Vietnam/ or Vietnam*.tw. or exp Yemen/ or Yemen*.tw. or exp Zambia/ or Zambia*.tw. or exp Zimbabwe/ or Zimbabwe*.tw. [mp=title, abstract, heading word, drug trade name, original title, device manufacturer, drug manufacturer, device trade name, keyword heading word, floating subheading word, candidate term word]	2590631

62	41 and 52 and 60 and 61	3398

| **Author** | **Year** | **Title** | **Country** | **Disaster/**  **Emergency** | **Population** | **Learning Method** | **Major topic** | **Study design** |
| --- | --- | --- | --- | --- | --- | --- | --- | --- |
| Panda R | 2022 | Evaluation of COVID-19 ECHO training program for healthcare workers in India - A Mixed-Method Study. | India | COVID-19 | Health workforce | Virtual training | Knowledge evaluation | Cross-sectional |
| Camputaro LA | 2021 | Intensive competency-based training strategy in a National Hospital in times of Pandemic. | El Salvador | COVID-19 | Health workforce | In-person training | Knowledge evaluation | Qualitative |
| Siddiqui SS; | 2023 | The impact of a "short-term" basic intensive care training program on the knowledge of nonintensivist doctors during the COVID-19 pandemic: An experience from a population-dense low- and middle-income country. | India | COVID-19 | Health workforce | In-person training | Knowledge evaluation | Observational |
| Jordan P | 2023 | Development of a training programme for professional nurses in South Africa - An educational response to the COVID-19 pandemic. | South Africa | COVID-19 | Health workforce | In-person training | Learning technique | Report |
| Kharel R | 2022 | Training program for female community volunteers to combat COVID 19 in rural Nepal. | Nepal | COVID-19 | Experts and volunteers | Virtual training | Learning technique | Report |
| Caviglia M | 2022 | Response to Mass-Casualty Incidents and Outbreaks: A Prehospital Disaster Training Package Developed for the National Emergency Medical Service in Sierra Leone. | Sierra Leone | Disasters | Health workforce | In-person training | Learning technique | Report |
| Singh SS | 2022 | Training community health workers for the COVID-19 response, India. | India | COVID-19 | Health workforce | In-person training | Impact | Cross-sectional |
| Guragai M | 2020 | Medical Students' Response to the COVID-19 Pandemic: Experience and Recommendations from Five Countries. | Brazil, Nepal, the Philippines, Rwanda, and the United States. | COVID-19 | Health workforce and community | Blended | Learning technique | Report |
| Marsh RH; | 2021 | Facing COVID-19 in Liberia: Adaptations of the Resilient and Responsive Health Systems Initiative. | Liberia | COVID-19 | Health workforce | Virtual training | Learning technique | Report |
| Uttekar S | 2023 | Empowering Health Workers to Build Public Trust in Vaccination: Experience from the International Pediatric Association's Online Vaccine Trust Course, 2020-2021. | International | COVID-19 | Health workforce | Virtual training | Knowledge evaluation | Report |
| Zerdo Z; | 2022 | Implementation of a malaria prevention education intervention in Southern Ethiopia: a qualitative evaluation. | Etihopia | others | Health workforce | In-person training | Impact | RCT |
| Perera N | 2022 | Implementation of a coronavirus disease 2019 infection prevention and control training program in a low-middle income country. | Syria | COVID-19 | Health workforce | In-person training | Knowledge evaluation | Report |
| Cuen AJ | 2022 | Fighting COVID-19 and HIV through community mobilisation: lessons from an integrated approach to the Africa CDC Partnership to Accelerate COVID-19 Testing (PACT) initiative in seven countries. | Africa | COVID-19 | Health workforce and community | Blended | Learning technique | Review |
| Malik JA; | 2022 | Myths and misconception of COVID-19 among hospital sanitary workers in Pakistan: Efficacy of a training program intervention. | Pakistan | COVID-19 | Health workforce | Virtual training | Knowledge evaluation | Before-after |
| Osula VO | 2022 | COVID-19 advanced respiratory care educational training programme for healthcare workers in Lesotho: an observational study. | Lesotho | COVID-19 | Health workforce | In-person training | Knowledge evaluation | Before-after |
| Cianelli R | 2013 | Mental health training experiences among Haitian healthcare workers post-earthquake 2010. | Haiti | Earthquake | Health workforce | In-person training | Impact | Observational |
| Lu Y | 2016 | Chinese military medical teams in the Ebola outbreak of Sierra Leone. | Sierra Leone | Ebola | Military | In-person training | Learning technique | Report |
| Yilmaz | 2021 | RE-AIMing COVID-19 online learning for medical students: a massive open online course evaluation. | Turkey | COVID-19 | Health workforce | Virtual training | Impact | Cross-sectional |
| Pek JH | 2020 | Teaching Disaster Site Medical Support in Indonesia. | Indonesia | Disasters | Health workforce | In-person training | Learning technique | Report |
| He LX | 2022 | Perspectives of nursing directors on emergency nurse deployment during the pandemic of COVID-19: A nationwide cross-sectional survey in mainland China | China | COVID-19 | Health workforce | N/A | Emergency plans | Cross-sectional |
| Ray S | 2021 | Innovation in primary health care responses to COVID-19 in Sub-Saharan Africa. | Africa | COVID-19 | Health workforce | N/A | Emergency plans | Review |
| Kochis M; | 2021 | Learning During and From a Crisis: The Student-Led Development of a COVID-19 Curriculum | International | COVID-19 | Health workforce | In-person training | Learning technique | Report |
| Shahrin L | 2022 | In-person training on COVID-19 case management and infection prevention and control: Evaluation of healthcare professionals in Bangladesh. | Bangladesh | COVID-19 | Health workforce | In-person training | Knowledge evaluation | Cross-sectional |
| Müller SA | 2020 | Implementation of the WHO hand hygiene strategy in Faranah regional hospital, Guinea. | Guinea | Ebola | Health workforce | In-person training | Knowledge evaluation | Before-after |
| Fredricks K; | 2017 | Community Health Workers and Disasters: Lessons Learned from the 2015 Earthquake in Nepal. | Nepal | Earthquake | Health workforce | N/A | Emergency plans | Qualitative |
| Eardley W | 2016 | Education and Ebola: initiating the cascade of emergency healthcare training. | Africa | Ebola | Health workforce | In-person training | Knowledge evaluation | Before-after |
| Chemali Z; | 2017 | Humanitarian space and well-being: effectiveness of training on a psychosocial intervention for host community-refugee interaction. | Lebanon | War | Experts and volunteers | In-person training | Impact | Before-after |
| Jaguga F | 2020 | Mental health response to the COVID-19 pandemic in Kenya: a review. | Kenya | COVID-19 | Health workforce and community | Virtual training | Emergency plans | Review |
| Ntahobakurira I; | 2011 | The Rwanda Field Epidemiology and Laboratory Training | Rwanda | others | Health workforce | In-person training | Learning technique | Report |
| Soeters HM | 2018 | Infection prevention and control training and capacity building during the Ebola epidemic in Guinea. | Guinea | Ebola | Health workforce | In-person training | Knowledge evaluation | Before-after |
| Stander M; | 2011 | Hospital disaster planning in the Western cape, South Africa. | South Africa | Disasters | Health workforce | N/A | Emergency plans | Cross-sectional |
| Naghavi Alhosseini | 2018 | Earthquake in the city: using real life gamification model for teaching professional commitment in high school students. | Iran | Earthquake | Citizens and affected population | Virtual training | Emergency plans | Before-after |
| Setiawan E | 2021 | Evaluating knowledge and skill in surgery clerkship during covid 19 pandemics: A single-center experience in Indonesia. | Indonesia | COVID-19 | Health workforce | In-person training | Knowledge evaluation | Cross-sectional |
| Dababnah S | 2019 | Feasibility of a trauma-informed parent-teacher cooperative training program for Syrian refugee children with autism. | Turkey | War | Citizens and affected population | In-person training | Impact | Before-after |
| Hawkes M; | 2009 | Use and limitations of malaria rapid diagnostic testing by community health workers in war-torn Democratic Republic of Congo. | Congo | others | Health workforce | In-person training | Knowledge evaluation | Before-after |
| Bustamante ND | 2020 | The Haiti Humanitarian Response Course: A Novel Approach to Local Responder Training in International Humanitarian Response. | Haiti | Disasters | Health workforce | Virtual training | Learning technique | Report |
| Wuthisuthimethawee P | 2022 | How the ARCH Project Could Contribute to Strengthening ASEAN Regional Capacities on Disaster Health Management (DHM). | ASEAN Member States | Disasters | Health workforce | Simulation | Learning technique | Report |
| Burlew R | 2014 | Assessing the relevance, efficiency, and sustainability of HIV/AIDS in-service training in Nigeria. | Nigeria | others | Health workforce | Virtual training | Emergency plans | Report |
| Thomas JJ; | 2022 | Participatory Workshop-Based Intervention for Better Preparedness and Awareness About Disaster Management Among Accredited Social Health Activists in India: A Brief Report. | India | Disasters | Health workforce | Simulation | Knowledge evaluation | Before-after |
| Joseph JK | 2012 | Lay health workers and HIV care in rural Lesotho: a report from the field. | Lesotho | others | Health workforce | In-person training | Knowledge evaluation | Cross-sectional |
| Patel U; | 2015 | Ebola Outbreak in Nigeria: Increasing Ebola Knowledge of Volunteer Health Advisors. | Nigeria | Ebola | Experts and volunteers | In-person training | Knowledge evaluation | Before-after |
| Çiçek A | 2020 | Combat medic course: evaluation of trainees' perception of learning and academic-self perception. | Turkey | War | Health workforce | In-person training | Impact | Before-after |
| Sonenthal PD | 2022 | Applying the WHO-ICRC BEC course to train emergency and inpatient healthcare workers in Sierra Leone early in the COVID-19 outbreak. | Sierra Leone | COVID-19 | Health workforce | In-person training | Knowledge evaluation | Before-after |
| Macfarlane C; | 2006 | Training of disaster managers at a masters degree level: from emergency care to managerial control. | South Africa | Disasters | Health workforce | Academic training | Learning technique | Report |
| Kang HM | 2021 | Development of a Medical Support Training Program for Disaster Management in Indonesia: A Hospital Disaster Medical Support Program for Indonesia. | Indonesia | Disasters | Health workforce | Simulation | Learning technique | Report |
| Magaña-Valladares L | 2018 | A MOOC as an immediate strategy to train health personnel in the cholera outbreak in Mexico. | Mexico | Cholera | Health workforce | Virtual training | Learning technique | Report |
| de Morais Pinto R | 2021 | Analyzing the reach of public health campaigns based on multidimensional aspects: the case of the syphilis epidemic in Brazil. | Brazil | others | Citizens and affected population | N/A | Emergency plans | Before-after |
| Engelbrecht B | 2021 | Prioritizing people and rapid learning in times of crisis: A virtual learning initiative to support health workers during the COVID-19 pandemic. | South Africa | COVID-19 | Health workforce | Virtual training | Learning technique | Report |
| Zhou M | 2020 | Research on the individualized short-term training model of nurses in emergency isolation wards during the outbreak of COVID-19. | China | COVID-19 | Health workforce | Blended | Impact | Before-after |
| Leitch L | 2009 | A case for using biologically-based mental health intervention in post-earthquake china: evaluation of training in the trauma resiliency model. | China | Earthquake | Health workforce | In-person training | Impact | Report |
| Ma D | 2021 | Does theme game-based teaching promote better learning about disaster nursing than scenario simulation: A randomized controlled trial. | China | Disasters | Health workforce | Virtual training | Knowledge evaluation | RCT |
| Lee PH | 2018 | The effectiveness of an on-line training program for improving knowledge of fire prevention and evacuation of healthcare workers: A randomized controlled trial. | China | Disasters | Health workforce | Virtual training | Emergency plans | RCT |
| Davidson PM | 2021 | Global digital social learning as a strategy to promote engagement in the era of COVID-19. | International | COVID-19 | Health workforce | Virtual training | Learning technique | Observational |
| Gul S | 2008 | Multitasking a telemedicine training unit in earthquake disaster response: paraplegic rehabilitation assessment. | Pakistan | Earthquake | Citizens and affected population | Virtual training | Learning technique | Before-after |
| Ng YM | 2020 | Coronavirus disease (COVID-19) prevention: Virtual classroom education for hand hygiene. | Hong Kong | COVID-19 | Health workforce | Virtual training | Learning technique | Report |
| Patel | 2022 | Simulation-based ventilatory training for the caregivers at primary and rural health care workers in Central India for dealing with COVID-19 pandemic: recommendations. | India | COVID-19 | Health workforce | Simulation | Learning technique | Report |
| Mutabaruka E | 2011 | The West Africa Field Epidemiology and Laboratory Training Program, a strategy to improve disease surveillance and epidemic control in West Africa. | Africa | others | Health workforce | In-person training | Learning technique | Opinion |
| Sommerland N | 2020 | Reducing HIV- and TB-Stigma among healthcare co-workers in South Africa: Results of a cluster randomised trial. | South Africa | others | Health workforce | In-person training | Knowledge evaluation | RCT |
| Furkan Dağcioğlu B | 2020 | Social adaptation status of Syrian refugee physicians living in Turkey. | Syria | War | Health workforce | N/A | Impact | Cross-sectional |
| Kuhls DA | 2017 | Basic Disaster Life Support (BDLS) Training Improves First Responder Confidence to Face Mass-Casualty Incidents in Thailand. | Thailand | Disasters | Health workforce | In-person training | Impact | Before-after |
| Feldman M | 2021 | Community health worker knowledge, attitudes and practices towards COVID-19: Learnings from an online cross-sectional survey using a digital health platform, UpSCALE, in Mozambique. | Mozambique | COVID-19 | Health workforce | Virtual training | Knowledge evaluation | Cross-sectional |
| Dunin-Bell O | 2018 | What do They Know? Guidelines and Knowledge Translation for Foreign Health Sector Workers Following Natural Disasters. | International | Disasters | Health workforce | N/A | Emergency plans | Review |
| Gunnlaugsson G | 2019 | Tiny Iceland' preparing for Ebola in a globalized world. | Iceland | Ebola | Health workforce | In-person training | Knowledge evaluation | Qualitative |
| El-Khani A | 2021 | Enhancing Teaching Recovery Techniques (TRT) with Parenting Skills: RCT of TRT + Parenting with Trauma-Affected Syrian Refugees in Lebanon Utilising Remote Training with Implications for Insecure Contexts and COVID-19. | Syria | War | Citizens and affected population | Virtual training | Impact | RCT |
| Liu L | 2012 | Zero Health Worker Infection: Experiences From the China Ebola Treatment Unit During the Ebola Epidemic in Liberia. | Liberia | Ebola | Health workforce | In-person training | Knowledge evaluation | Report |
| James LE | 2020 | Integrating mental health and disaster preparedness in intervention: a randomized controlled trial with earthquake and flood-affected communities in Haiti. | Haiti | Earthquake | Citizens and affected population | In-person training | Impact | RCT |
| Chua | 2008 | Building partnerships to address the HIV epidemic. | Singapore | others | Health workforce | In-person training | Impact | Report |
| Cruz-Vega | 2016 | [Experience in training in emergencies, Division of Special Projects in Health, Instituto Mexicano del Seguro Social]. | Mexico | Disasters | Health workforce and community | Blended | Emergency plans | Report |
| Van Heng | 2008 | Non-doctors as trauma surgeons? A controlled study of trauma training for non-graduate surgeons in rural Cambodia. | Cambodia | War | Experts and volunteers | In-person training | Emergency plans | Before-after |
| Talisuna AO | 2020 | The COVID-19 pandemic: broad partnerships for the rapid scale up of innovative virtual approaches for capacity building and credible information dissemination in Africa. | Africa | COVID-19 | Health workforce | Virtual training | Learning technique | Report |
| Ren | 2017 | Experiences in disaster-related mental health relief work: An exploratory model for the interprofessional training of psychological relief workers. | China | Earthquake | Health workforce | In-person training | Impact | Qualitative |
| Najafi Ghezeljeh T; | 2019 | Effect of education using the virtual social network on the knowledge and attitude of emergency nurses of disaster preparedness: A quasi-experiment study. | Iran | Disasters | Health workforce | Virtual training | Knowledge evaluation | Before-after |
| Hou | 2018 | Disaster Medicine in China: Present and Future. | China | Disasters | Health workforce | In-person training | Emergency plans | Report |
| McQuilkin | 2017 | Academic Medical Support to the Ebola Virus Disease Outbreak in Liberia. | Africa | Ebola | Health workforce | In-person training | Emergency plans | Report |
| Hemingway-Foday JJ; | 2020 | Lessons Learned from Reinforcing Epidemiologic Surveillance During the 2017 Ebola Outbreak in the Likati District, Democratic Republic of the Congo. | Congo | Ebola | Health workforce | In-person training | Emergency plans | Report |
| Yao K | 2010 | Ensuring the quality of HIV rapid testing in resource-poor countries using a systematic approach to training. | Africa | others | Health workforce | In-person training | Learning technique | Report |
| Bodas | 2022 | Training Package for Emergency Medical Teams Deployed to Disaster Stricken Areas: Has 'TEAMS' Achieved its Goals? | Italy | Disasters | Health workforce | In-person training | Knowledge evaluation | Before-after |
| Bemah P | 2019 | Strengthening healthcare workforce capacity during and post Ebola outbreaks in Liberia: an innovative and effective approach to epidemic preparedness and response. | Liberia | Ebola | Health workforce | In-person training | Knowledge evaluation | Report |
| Oji MO | 2018 | Implementing infection prevention and control capacity building strategies within the context of Ebola outbreak in a "Hard-to-Reach" area of Liberia. | Liberia | Ebola | Health workforce | In-person training | Knowledge evaluation | Report |
| Najarian | 2004 | Disaster intervention: long-term psychosocial benefits in Armenia | Armenia | Earthquake | Health workforce | In-person training | Emergency plans | Opinion |
| Wang | 2021 | The effectiveness of E-learning in continuing medical education for tuberculosis health workers: a quasi-experiment from China. | China | others | Health workforce | Virtual training | Knowledge evaluation | Before-after |
| Limpakarnjanarat | 2007 | Long-term capacity-building in public health emergency preparedness in Thailand--short report. | Thailand | Disasters | Health workforce | N/A | Emergency plans | Report |
| Djalali | 2009 | A fundamental, national, medical disaster management plan: an education-based model. | Iran | Earthquake | Health workforce | In-person training | Knowledge evaluation | Before-after |
| Carlos | 2015 | Hospital preparedness for Ebola virus disease: a training course in the Philippines. | Philippines | Ebola | Health workforce | In-person training | Knowledge evaluation | Before-after |
| Cherian | 2004 | Essential emergency surgical, procedures in resource-limited facilities: a WHO workshop in Mongolia. | China | Disasters | Health workforce | Blended | Learning technique | Report |
| Welton-Mitchell | 2018 | An integrated approach to mental health and disaster preparedness: a cluster comparison with earthquake affected communities in Nepal. | Nepal | Earthquake | Citizens and affected population | N/A | Impact | Before-after |
| Wang | 2010 | Improving emergency preparedness capability of rural public health personnel in China. | China | Disasters | Health workforce | In-person training | Knowledge evaluation | Before-after |
| Vijaykumar | 2006 | Psychosocial interventions after tsunami in Tamil Nadu, India. | India | Disasters | Experts and volunteers | N/A | Impact | Report |
| Koca | 2020 | The effect of the disaster management training program among nursing students. | Turkey | Disasters | Health workforce | In-person training | Impact | RCT |
| Zhang | 2021 | Effect of virtual reality simulation training on the response capability of public health emergency reserve nurses in China: a quasiexperimental study. | China | COVID-19 | Health workforce | Virtual training | Knowledge evaluation | RCT |
| Rosa | 2021 | A Virtual Coaching Workshop for a Nurse-Led Community-Based Palliative Care Team in Liberia, West Africa, to Promote Staff Well-Being During COVID-19. | Liberia | COVID-19 | Health workforce | Virtual training | Impact | Before-after |
| Rajasingham | 2011 | Cholera prevention training materials for community health workers, Haiti, 2010–2011. | Haiti | Cholera | Health workforce | In-person training | Learning technique | Report |
| Shin YA | 2018 | The Effectiveness of International Non-Governmental Organizations' Response Operations during Public Health Emergency: Lessons Learned from the 2014 Ebola Outbreak in Sierra Leone. | Sierra Leone | Ebola | Health workforce and community | N/A | Learning technique | Report |
| Ripp JA | 2012 | The response of academic medical centers to the 2010 Haiti earthquake: the Mount Sinai School of Medicine experience. | Haiti | Earthquake | Health workforce | In-person training | Knowledge evaluation | Report |
| Maduka | 2015 | Ethical challenges of containing Ebola: the Nigerian experience. | Nigeria | Ebola | Health workforce | In-person training | Emergency plans | Report |
| Olu O | 2018 | What should the African health workforce know about disasters? Proposed competencies for strengthening public health disaster risk management education in Africa. | Africa | Disasters | Health workforce | Academic training | Emergency plans | Review |
| Math | 2006 | Tsunami: psychosocial aspects of Andaman and Nicobar islands. Assessments and intervention in the early phase. | India | Disasters | Health workforce | In-person training | Emergency plans | Report |
| Chamane | 2022 | The effect of a mobile-learning curriculum on improving compliance to quality management guidelines for HIV rapid testing services in rural primary healthcare clinics, KwaZulu-Natal, South Africa: a quasi-experimental study. | South Africa | others | Health workforce | Virtual training | Knowledge evaluation | Before-after |
| Bazeyo | 2013 | Regional approach to building operational level capacity for disaster planning: the case of the Eastern Africa region. | Africa | Disasters | Health workforce | N/A | Emergency plans | Report |
| Yi | 2018 | Developing and implementing a global emergency medicine course: Lessons learned from Rwanda. | Rwanda | Disasters | Health workforce | In-person training | Learning technique | Report |
| Hébert | 2020 | Video as a public health knowledge transfer tool in Burkina Faso: A mixed evaluation comparing three narrative genres. | Burquina Faso | others | Health workforce | Virtual training | Learning technique | Before-after |
| Orach | 2013 | Use of the Automated Disaster and Emergency Planning Tool in developing district level public health emergency operating procedures in three East African countries. | Africa | Disasters | National Institutions | Virtual training | Emergency plans | Report |
| Bazeyo | 2015 | Ebola a reality of modern Public Health; need for Surveillance, Preparedness and Response Training for Health Workers and other multidisciplinary teams: a case for Uganda. | Uganda | Ebola | Health workforce and community | Blended | Emergency plans | Report |
| Sharara-Chami | 2020 | In Situ Simulation: An Essential Tool for Safe Preparedness for the COVID-19 Pandemic | Lebanon | COVID-19 | Health workforce | Simulation | Impact | Before-after |
| El-Bahnasawy | 2014 | Selected infectious disease disasters for nursing staff training at Egyptian Eastern Border | Egypt | others | Health workforce | N/A | Knowledge evaluation | Before-after |
| Leow | 2012 | Mass casualty incident training in a resource-limited environment. | Sierra Leone | Disasters | Health workforce | In-person training | Knowledge evaluation | Before-after |
| Olness | 2005 | Training of health care professionals on the special needs of children in the management of disasters: experience in Asia, Africa, and Latin America. | International | Disasters | Health workforce | In-person training | Impact | Report |
| Otu | 2016 | Using a mHealth tutorial application to change knowledge and attitude of frontline health workers to Ebola virus disease in Nigeria: a before-and-after study | Nigeria | Ebola | Health workforce | Virtual training | Knowledge evaluation | Before-after |
| Orach | 2013 | Performance of district disaster management teams after undergoing an operational level planners' training in Uganda. | Uganda | Disasters | Health workforce | N/A | Emergency plans | Report |
| Pérez-Manchón | 2015 | [Telemedicine, a medical social network for humanitarian aid between Spain and Cameroon]. | Cameroon | Disasters | Health workforce | Virtual training | Learning technique | Report |
| Meade | 2007 | A deployable telemedicine capability in support of humanitarian operations. | Africa | Disasters | Health workforce | Virtual training | Learning technique | Report |
| Findyartini | 2021 | Supporting newly graduated medical doctors in managing COVID-19: An evaluation of a Massive Open Online Course in a limited-resource setting. | Indonesia | COVID-19 | Health workforce | Virtual training | Impact | Before-after |
| Hess | 2004 | Development of emergency medical services in Guatemala. | Guatemala | Disasters | Experts and volunteers | N/A | Emergency plans | Report |
| Yamada | 2007 | Interdisciplinary problem-based learning as a method to prepare Micronesia for public health emergencies. | Hawaii | Disasters | Health workforce | N/A | Emergency plans | Report |
| Kizakevich | 2007 | Virtual simulation-enhanced triage training for Iraqi medical personnel. | Iraq | Disasters | Health workforce | In-person training | Learning technique | Report |
| O'Reilly G | 2008 | In the wake of Sri Lanka's tsunami: the health for the south capacity-building project. | Sri Lanka | Disasters | Health workforce | N/A | Emergency plans | Report |
| Sullivan J | 2021 | The Impact of Simulation-Based Education on Nurses' Perceived Predeployment Anxiety During the COVID-19 Pandemic Within the Cultural Context of a Middle Eastern Country. | Qatar | COVID-19 | Health workforce | Simulation | Impact | Before-after |
| Tegegne MD | 2022 | Use of social media for COVID-19-related information and associated factors among health professionals in Northwest Ethiopia: A cross-sectional study. | Ethiopia | COVID-19 | Health workforce | Virtual training | Knowledge evaluation | Cross-sectional |
| Jafree | 2022 | WhatsApp-Delivered Intervention for Continued Learning for Nurses in Pakistan During the COVID-19 Pandemic: Results of a Randomized-Controlled Trial. | Pakistan | COVID-19 | Health workforce | Virtual training | Knowledge evaluation | RCT |
| Leichner A; | 2021 | Mental health integration in primary health services after the earthquake in Nepal: a mixed-methods program evaluation. | Nepal | Earthquake | Health workforce and community | In-person training | Knowledge evaluation | Cross-sectional |
| Ng | 2009 | China-Australia training on psychosocial crisis intervention: response to the earthquake disaster in Sichuan. | China | Earthquake | Health workforce | In-person training | Impact | Before-after |
| AlAssaf | 2022 | Challenges in Pandemic Disaster Preparedness: Experience of a Saudi Academic Medical Center. | Saudi Arabia | COVID-19 | Health workforce | Blended | Emergency plans | Report |
| Button GJ | 2022 | Utilizing a "Crawl, Walk, Run" Training Model to Enhance Field Sanitation Capabilities for Peacekeeping Forces: A Recommendation for the Department of Defense Global Health Engagement Enterprise. | Senegal | COVID-19 | Military | Mixed | Learning technique | Report |
| Oliveira | 2020 | Personal Protective Equipment in the coronavirus pandemic: training with Rapid Cycle Deliberate Practice. | Brazil | COVID-19 | Health workforce | Simulation | Knowledge evaluation | Report |
| Morton Hamer MJ | 2019 | Enhancing Global Health Security: US Africa Command's Disaster Preparedness Program. | Africa | Disasters | Health workforce | In-person training | Emergency plans | Report |
| Khan JA | 2020 | Impact of multi-professional simulation-based training on perceptions of safety and preparedness among health workers caring for coronavirus disease 2019 patients in Pakistan. | Pakistan | COVID-19 | Health workforce | Simulation | Knowledge evaluation | Before-after |
| Jordans MJ | 2012 | Evaluation of a brief training on mental health and psychosocial support in emergencies: a pre- and post-assessment in Nepal. | Nepal | disasters | Health workforce | In-person training | Knowledge evaluation | Before-after |
| Lubogo M | 2015 | Ebola virus disease outbreak; the role of field epidemiology training programme in the fight against the epidemic, Liberia, 2014. | Liberia | Ebola | Health workforce | In-person training | Learning technique | Report |
| Lin L; | 2014 | The public health system response to the 2008 Sichuan province earthquake: a literature review and interviews. | China | Earthquake | National Institutions | N/A | Emergency plans | Report |
| Xia | 2020 | Evaluating the effectiveness of a disaster preparedness nursing education program in Chengdu, China. | China | Disasters | Health workforce | In-person training | Knowledge evaluation | RCT |
| Bajow N; | 2022 | Assessment of the effectiveness of a course in major chemical incidents for front line health care providers: a pilot study from Saudi Arabia. | Saudi Arabia | others | Health workforce | Simulation | Learning technique | Before-after |
| Saghafinia M | 2009 | Effect of the rural rescue system on reducing the mortality rate of landmine victims: a prospective study in Ilam Province, Iran. | Iran | others | Health workforce | In-person training | Knowledge evaluation | Observational |
| He | 2021 | Practice in Information Technology Support for Fangcang Shelter Hospital during COVID-19 Epidemic in Wuhan, China. | China | COVID-19 | Health workforce | Virtual training | Impact | Report |
| Kenar | 2006 | Medical preparedness against chemical and biological incidents for the NATO Summit in Istanbul and lessons learned. | Turkey | Disasters | Health workforce | In-person training | Emergency plans | Report |
| Salita C | 2019 | Development, implementation, and evaluation of a lay responder disaster training package among school teachers in Angeles City, Philippines: using Witte's behavioral model. | Philippines | Disasters | Experts and volunteers | In-person training | Knowledge evaluation | Before-after |
| Tauxe RV; | 2011 | Rapid development and use of a nationwide training program for cholera management, Haiti, 2010. | Haiti | Cholera | Health workforce | In-person training | Learning technique | Before-after |
| Werdhani RA | 2022 | A COVID-19 self-isolation monitoring module for FMUI undergraduate medical students: Linking learning and service needs during the pandemic surge in Indonesia. | Indonesia | COVID-19 | Health workforce | Virtual training | Impact | Report |
| Alshiekhly U | 2015 | Facebook as a learning environment for teaching medical emergencies in dental practice. | Syria | others | Health workforce | Virtual training | Impact | Cross-sectional |
| Cai W; | 2022 | Doctor of Public Health-Crisis Management and COVID-19 Prevention and Control: A Case Study in China. | China | COVID-19 | Health workforce | In-person training | Emergency plans | Report |
| Hageman | 2016 | Infection Prevention and Control for Ebola in Health Care Settings - West Africa and United States. | Africa | Ebola | Health workforce | In-person training | Emergency plans | Report |
| Pang | 2009 | Pilot training program for developing disaster nursing competencies among undergraduate students in China. | China | Disasters | Health workforce | In-person training | Knowledge evaluation | Before-after |
| Brisebois | 2011 | The Role 3 Multinational Medical Unit at Kandahar Airfield 2005-2010. | Afghanistan | War | Health workforce | Simulation | Emergency plans | Report |
| Gertler M | 2018 | West Africa Ebola outbreak - immediate and hands-on formation: the pre-deployment training program for frontline aid workers of the German Red Cross, other aid organizations, and the German Armed Forces, Wuerzburg, Germany 2014/15 | Africa | Ebola | Health workforce | In-person training | Impact | Report |
| Alim S | 2015 | Evaluation of disaster preparedness training and disaster drill for nursing students. | Indonesia | Disasters | Health workforce | In-person training | Knowledge evaluation | Before-after |
| Boon | 2009 | The impact of a community-based pilot health education intervention for older people as caregivers of orphaned and sick children as a result of HIV and AIDS in South Africa. | South Africa | others | Citizens and affected population | In-person training | Impact | Report |
| Subedi | 2018 | The Health Sector Response to the 2015 Earthquake in Nepal. | Nepal | Earthquake | Health workforce | In-person training | Emergency plans | Report |
| Cooper | 2012 | Evaluating the efficacy of the AAP "pediatrics in disaster" course: the Chinese experience. | International | Disasters | Health workforce | N/A | Impact | Before-after |
| Evans | 2016 | Innovation in Graduate Education for Health Professionals in Humanitarian Emergencies. | International | Disasters | Academia | In-person training | Learning technique | Report |
| Silva | 2021 | Implementation of COVID-19 telemonitoring: repercussions in Nursing academic training. | Brazil | COVID-19 | Health workforce | Virtual training | Knowledge evaluation | Report |
| Aghababaeian | 2013 | A comparative study of the effect of triage training by role-playing and educational video on the knowledge and performance of emergency medical service staffs in Iran. | Iran | Disasters | Health workforce | Blended | Knowledge evaluation | RCT |
| Chiu | 2021 | Facing the Coronavirus Pandemic: An Integrated Continuing Education Program in Taiwan. | China | COVID-19 | Health workforce | Virtual training | Knowledge evaluation | Before-after |
| Haar | 2020 | Strong families: a new family skills training programme for challenged and humanitarian settings: a single-arm intervention tested in Afghanistan. | Afghanistan | Disasters | Citizens and affected population | In-person training | Impact | Before-after |
| Shi | 2020 | A simulation training course for family medicine residents in China managing COVID-19. | China | COVID-19 | Health workforce | Simulation | Knowledge evaluation | Before-after |
| Rouzier | 2013 | Cholera vaccination in urban Haiti. | Haiti | Cholera | Health workforce and community | In-person training | Emergency plans | Report |
| Abbas | 2018 | Peers versus professional training of basic life support in Syria: a randomized controlled trial. | Syria | Disasters | Health workforce | In-person training | Impact | RCT |
| Finnegan | 2015 | Preparing British Military nurses to deliver nursing care on deployment. An Afghanistan study. | Afghanistan | Disasters | Military | In-person training | Emergency plans | Report |
| Schreiber | 2004 | Hospital preparedness for possible nonconventional casualties: an Israeli experience. | Israel | others | Health workforce | In-person training | Emergency plans | Report |
| Phillips GA | 2014 | Capacity building for emergency care: Training the first emergency specialists in Myanmar. | Myanmar | Disasters | Health workforce | Academic training | Learning technique | Report |
| Sun L | 2021 | Intervention Effect of Time Management Training on Nurses' Mental Health during the COVID-19 Epidemic. | China | COVID-19 | Health workforce | In-person training | Impact | Before-after |
| Hung KKC | 2021 | Health Workforce Development in Health Emergency and Disaster Risk Management: The Need for Evidence-Based Recommendations. | International | Disasters | Health workforce | N/A | Emergency plans | Review |
| Mosquera A | 2015 | U.S. Public Health Service Response to the 2014-2015 Ebola Epidemic in West Africa: A Nursing Perspective. | Africa | Ebola | Health workforce | Simulation | Learning technique | Report |
| Bajow NA | 2019 | A Basic Course in Humanitarian Health Emergency and Relief: A Pilot Study from Saudi Arabia. | Saudi Arabia | Disasters | Health workforce | In-person training | Knowledge evaluation | Before-after |
| Chan SS | 2010 | Development and evaluation of an undergraduate training course for developing International Council of Nurses disaster nursing competencies in China. | China | Disasters | Health workforce | Academic training | Knowledge evaluation | Before-after |
| Ikeda S | 2022 | Introduction to the Project for Strengthening the ASEAN Regional Capacity on Disaster Health Management (ARCH Project). | Japan | Disasters | Health workforce | Academic training | Emergency plans | Report |
| Iskanderani AI | 2021 | Artificial Intelligence and Medical Internet of Things Framework for Diagnosis of Coronavirus Suspected Cases. | China | COVID-19 | Health workforce | Virtual training | Learning technique | Report |
| Rehman H | 2020 | Effectiveness of basic training session regarding the awareness of Ebola virus disease among nurses of public tertiary care hospitals of Lahore. | Pakistan | Ebola | Health workforce | In-person training | Knowledge evaluation | Before-after |
| Díaz-Guio DA | 2020 | Cognitive load and performance of health care professionals in donning and doffing PPE before and after a simulation-based educational intervention and its implications during the COVID-19 pandemic for biosafety. | Colombia | COVID-19 | Health workforce | Simulation | Knowledge evaluation | Before-after |
| Bai HX | 2020 | Artificial Intelligence Augmentation of Radiologist Performance in Distinguishing COVID-19 from Pneumonia of Other Origin at Chest CT. | China | COVID-19 | Health workforce | Virtual training | Knowledge evaluation | Observational |
| Tan W; | 2020 | Whole-Process Emergency Training of Personal Protective Equipment Helps Healthcare Workers Against COVID-19: Design and Effect. | China | COVID-19 | Health workforce | Simulation | Knowledge evaluation | Before-after |
| Kimani D | 2022 | Adopting World Health Organization Multimodal Infection Prevention and Control Strategies to Respond to COVID-19, Kenya. | Kenya | COVID-19 | Health workforce | In-person training | Learning technique | Report |
| Hu X; | 2022 | Creation and application of war trauma treatment simulation software for first aid on the battlefield based on undeformed high-resolution sectional anatomical image (Chinese Visible Human dataset). | China | Disasters | Health workforce | Virtual training | Impact | Report |
| Bajow N | 2015 | Proposal for a community-based disaster management curriculum for medical school undergraduates in Saudi Arabia | Saudi Arabia | Disasters | Health workforce | Academic training | Learning technique | Report |
| Kesavadev J; | 2021 | A new interventional home care model for COVID management: Virtual Covid IP. | India | COVID-19 | Health workforce | Virtual training | Learning technique | Observational |
| Sohn VY; | 2007 | From the combat medic to the forward surgical team: the Madigan model for improving trauma readiness of brigade combat teams fighting the Global War on Terror. | Iraq | War | Military | Simulation | Knowledge evaluation | Report |
| Cerqueira-Silva T | 2021 | Bridging Learning in Medicine and Citizenship During the COVID-19 Pandemic: A Telehealth-Based Case Study. | Brazil | COVID-19 | Health workforce | Virtual training | Impact | Report |
| Barbier O | 2018 | Has Current French Training for Military Orthopedic Surgeons Deployed in External Operations Been Appropriately Adapted? | Africa and Afghanistan | War | Military | Academic training | Learning technique | Observational |
| Leochico CFD; | 2021 | Role of Telerehabilitation in the Rehabilitation Medicine Training Program of a COVID-19 Referral Center in a Developing Country. | Philippines | COVID-19 | Health workforce | Virtual training | Learning technique | Report |
| Pereira BM | 2010 | Predeployment mass casualty and clinical trauma training for US Army forward surgical teams. | Iraq and Afghanistan | war | Military | In-person training | Impact | Report |
| Philip S | 2022 | A report on successful introduction of tele mental health training for primary care doctors during the COVID 19 pandemic. | India | COVID-19 | Health workforce | Virtual training | Knowledge evaluation | Report |
| Brearley MB | 2016 | Pre-deployment Heat Acclimatization Guidelines for Disaster Responders. | Philippines | Disasters | Military | Virtual training | Knowledge evaluation | Report |
| Das A; | 2022 | Implementation of infection prevention and control practices in an upcoming COVID-19 hospital in India: An opportunity not missed. | India | COVID-19 | Health workforce | Blended | Knowledge evaluation | Observational |
| Gareev I; | 2021 | The opportunities and challenges of telemedicine during COVID-19 pandemic. | International | COVID-19 | Health workforce | Virtual training | Learning technique | Report |
| Jensen | 2015 | Integration of Surgical Residency Training With US Military Humanitarian Missions. | South Asia | War | Military | In-person training | Knowledge evaluation | Report |
| Choufani | 2021 | Evaluation of a fellowship abroad as part of the initial training of the French military surgeon. | Africa | War | Military | In-person training | Impact | Report |
| Tashkandi | 2021 | Nursing strategic pillars to enhance nursing preparedness and response to COVID-19 pandemic at a tertiary care hospital in Saudi Arabia | Saudi Arabia | COVID-19 | Health workforce | In-person training | Knowledge evaluation | Report |
| Chiu | 2021 | Developing and Implementing a Dedicated Prone Positioning Team for Mechanically Ventilated ARDS Patients During the COVID-19 Crisis. | China | COVID-19 | Health workforce | In-person training | Knowledge evaluation | Report |
| Operario | 2016 | Effect of a knowledge-based and skills-based programme for physicians on risk of sexually transmitted reinfections among high-risk patients in China: a cluster randomised trial. | China | others | Health workforce | In-person training | Knowledge evaluation | RCT |
| Wang | 2009 | Intervention to train physicians in rural China on HIV/STI knowledge and risk reduction counseling: preliminary findings. | China | others | Health workforce | In-person training | Knowledge evaluation | Before-after |
| El-Bahnasawy | 2015 | TRAINING PROGRAM FOR NURSING STAFF REGARDING VIRAL HEMORRHAGIC FEVERS IN A MILITARY HOSPITAL. | Egypt | others | Health workforce | In-person training | Impact | Before-after |
| Khari | 2022 | The Effect of E-Learning Program for COVID-19 Patient Care on the Knowledge of Nursing Students: A Quasi-Experimental Study. | Iran | COVID-19 | Health workforce | Virtual training | Knowledge evaluation | Before-after |
| Ripoll-Gallardo | 2020 | Residents working with Médecins Sans Frontières: training and pilot evaluation. | Africa | Disasters | Health workforce | Blended | Impact | Report |
| Otu | 2021 | Training health workers at scale in Nigeria to fight COVID-19 using the InStrat COVID-19 tutorial app: an e-health interventional study. | Nigeria | COVID-19 | Health workforce | Virtual training | Knowledge evaluation | Before-after |
| Lopes | 2020 | Adult learning and education as a tool to contain pandemics: The COVID-19 experience. | Africa | COVID-19 | Citizens and affected population | In-person training | Emergency plans | Opinion |
| Jackson | 2022 | Developing and Implementing Noninvasive Ventilator Training in Haiti during the COVID-19 Pandemic. | Haiti | COVID-19 | Health workforce | In-person training | Knowledge evaluation | Before-after |
| Sharma | 2021 | Effectiveness of Video-Based Online Training for Health Care Workers to Prevent COVID-19 Infection: An Experience at a Tertiary Care Level Institute, Uttarakhand, India. | India | COVID-19 | Health workforce | Virtual training | Knowledge evaluation | Before-after |
| Daniel | 2020 | Responding to Palliative Care Training Needs in the Coronavirus Disease 2019 Era: The Context and Process of Developing and Disseminating Training Resources and Guidance for Low- and Middle-Income Countries from Kerala, South India. | India | COVID-19 | Health workforce | Virtual training | Emergency plans | Report |
| Downie | 2022 | Remote Consulting in Primary Health Care in Low- and Middle-Income Countries: Feasibility Study of an Online Training Program to Support Care Delivery During the COVID-19 Pandemic. | Tanzania | COVID-19 | Health workforce | Virtual training | Knowledge evaluation | Report |
| Scott | 2020 | Training the Addiction Treatment Workforce in HIV Endemic Regions: An Overview of the South Africa HIV Addiction Technology Transfer Center Initiative. | South Africa | others | Experts and volunteers | In-person training | Knowledge evaluation | Before-after |
| Usami | 2018 | Addressing challenges in children's mental health in disaster-affected areas in Japan and the Philippines - highlights of the training program by the National Center for Global Health and Medicine. | Japan | Earthquake | Health workforce | In-person training | Impact | Report |
| Buyego | 2021 | Feasibility of Virtual Reality based Training for Optimising COVID-19 Case Handling in Uganda. | Uganda | COVID-19 | Health workforce | Virtual training | Impact | Report |
| Babu | 2021 | Simulated Patient Environment: A Training Tool for Healthcare Professionals in COVID-19 Era. | India | COVID-19 | Health workforce | Simulation | Knowledge evaluation | Before-after |
| Liu | 2022 | Development and Evaluation of Innovative and Practical Table-top Exercises Based on a Real Mass-Casualty Incident. | China | Disasters | Health workforce | Virtual training | Impact | Before-after |
| Fuenfer | 2009 | The U.S. military wartime pediatric trauma mission: how surgeons and pediatricians are adapting the system to address the need. | Afganistan and Iraq | War | Health workforce | Virtual training | Emergency plans | Report |
| Macht | 2022 | COVID-19: Development and implementation of a video-conference-based educational concept to improve the hygiene skills of health and nursing professionals in the Republic of Kosovo. | Kosovo | COVID-19 | Health workforce | Virtual training | Knowledge evaluation | Report |
| Jobson | 2019 | Targeted mentoring for human immunodeficiency virus programme support in South Africa | South Africa | others | Health workforce | In-person training | Impact | Report |
| Zelnick | 2018 | Training social workers to enhance patient-centered care for drug-resistant TB-HIV in South Africa. | South Africa | others | Health workforce | In-person training | Knowledge evaluation | Report |
| Cena-Navarro | 2022 | Biosafety Capacity Building During the COVID-19 Pandemic: Results, Insights, and Lessons Learned from an Online Approach in the Philippines. | Philippines | COVID-19 | Health workforce | Virtual training | Learning technique | Report |
| Richard | 2009 | Essential trauma management training: addressing service delivery needs in active conflict zones in eastern Myanmar. | Myanmar | War | Health workforce | In-person training | Knowledge evaluation | Report |
| Galagan | 2017 | Improving Tuberculosis (TB) and Human Immunodeficiency Virus (HIV) Treatment Monitoring in South Africa: Evaluation of an Advanced TB/HIV Course for Healthcare Workers. | South Africa | others | Health workforce | In-person training | Knowledge evaluation | Before-after |
| Irizarry | 2012 | Advanced Medical Technology Capacity Building and the Medical Mentoring Event: A Unique Application of SOF Counterinsurgency Medical Engagement Strategies. | Afghanistan | War | Health workforce and community | Simulation | Emergency plans | Report |
| Wanjiku | 2022 | Feasibility of project ECHO telementoring to build capacity among non-specialist emergency care providers. | Kenya | Disasters | Health workforce | Virtual training | Emergency plans | Report |
| Arnold | 2020 | Bridging the Gap Between Emergency Response and Health Systems Strengthening: The Role of Improvement Teams in Integrating Zika Counseling in Family Planning Services in Honduras. | Honduras | others | Health workforce | In-person training | Emergency plans | Before-after |
| Martinez | 2018 | Tourniquet Training Program Assessed by a New Performance Score. | Africa | War | Health workforce | In-person training | Knowledge evaluation | RCT |
| Kulshreshtha P | 2022 | Preparedness of Undergraduate Medical Students to Combat COVID-19: A Tertiary Care Experience on the Effectiveness and Efficiency of a Training Program and Future Prospects. | India | COVID-19 | Health workforce | Simulation | Knowledge evaluation | Before-after |
| Ahluwalia | 2021 | Effectiveness of remote practical boards and telesimulation for the evaluation of emergency medicine trainees in India. | India | Disasters | Health workforce | Virtual training | Impact | Report |
| Al-Hadidi | 2021 | Homemade cardiac and vein cannulation ultrasound phantoms for trauma management training in resource-limited settings. | Syria | Disasters | Health workforce | Simulation | Impact | Report |
| Patel | 2020 | "Emerging Technologies and Medical Countermeasures to Chemical, Biological, Radiological, and Nuclear (CBRN) Agents in East Ukraine" | Ukraine | War | Health workforce | In-person training | Emergency plans | Report |
| Wang | 2022 | Practical COVID-19 Prevention Training for Obstetrics and Gynecology Residents Based on the Conceive-Design-Implement-Operate Framework. | China | COVID-19 | Health workforce | Academic training | Impact | RCT |
| de Lesquen | 2020 | Adding the Capacity for an Intensive Care Unit Dedicated to COVID 19, Preserving the Operational Capability of a French Golden Hour Offset Surgical Team in Sahel. | Niger and Mali | COVID-19 | Military | Academic training | Emergency plans | Report |
| Ye | 2021 | Point-of-care training program on COVID-19 infection prevention and control for pediatric healthcare workers: a multicenter, cross-sectional questionnaire survey in Shanghai, China. | China | COVID-19 | Health workforce | Virtual training | Impact | Before-after |
| Bhattacharya | 2020 | Impact of a training program on disaster preparedness among paramedic students of a tertiary care hospital of North India: A single-group, before-after intervention study. | India | Disasters | Health workforce | Blended | Impact | Before-after |
| Wong | 2022 | ECMO simulation training during a worldwide pandemic: The role of ECMO telesimulation. | China | COVID-19 | Health workforce | Virtual simulation | Impact | Before-after |
| Fernández-Miranda | 2021 | Developing a Training Web Application for Improving the COVID-19 Diagnostic Accuracy on Chest X-ray. | Chile | COVID-19 | Health workforce | Virtual training | Knowledge evaluation | Report |
| Khoshnudi | 2022 | Comparison of the effect of bioterrorism education through two methods of lecture and booklet on the knowledge and attitude of nurses of Shams Al-Shomus Nezaja Hospital. | Iran | Disasters | Health workforce | In-person training | Knowledge evaluation | Before-after |
| Liesveld | 2022 | Teaching disaster preparedness to pre-licensure students: A collaborative project during the pandemic. | International | COVID-19 | Health workforce | Virtual training | Impact | Report |
| Shilkofski | 2017 | Pediatric Emergency Care in Disaster-Affected Areas: A Firsthand Perspective after Typhoons Bopha and Haiyan in the Philippines. | Philippines | Disasters | Health workforce | N/A | Emergency plans | Review |
| Bankole | 2021 | KNOWLEDGE OF HEALTH WORKERS ON CHOLERA MANAGEMENT IN OYO STATE: RESULTS OF A TRAINING INTERVENTION. | Nigeria | Cholera | Health workforce | In-person training | Knowledge evaluation | Before-after |
|  | 2023 | The Use of Open-Source Online Course Content for Training in Public Health Emergencies: Mixed Methods Case Study of a COVID-19 Course Series for Health Professionals | International | COVID-19 | Health workforce | Virtual training | Learning technique | Qualitative |
| Klomp | 2020 | CDC's Multiple Approaches to Safeguard the Health, Safety, and Resilience of Ebola Responders | Africa | Ebola | Health workforce | In-person training | Emergency plans | Report |
| Van Hulle | 2020 | Tailoring malaria routine activities within the covid-19 pandemic: A risk and mitigation assessment of eight countries in West Africa | West Africa | others | Health workforce and community | Virtual training | Emergency plans | Report |
| Guerrero-Torres | 2020 | Impact of Training Residents to Improve HIV Screening in a Teaching Hospital in Mexico City | Mexico | others | Health workforce | In-person training | Knowledge evaluation | Before-after |
| Sena | 2020 | Disaster preparedness training in emergency medicine residents using a tabletop exercise | International | Disasters | Health workforce | In-person training | Learning technique | Before-after |
| Mitchell | 2020 | A partnership to develop disaster simulations for nursing students in response to climate change: description of a programme in Bluefields, Nicaragua, and Virginia, USA | Nicaragua y USA | Disasters | Health workforce | In-person training | Learning technique | Report |
| Amini | 2019 | Epidemiological profile of crimean congo hemorrhagic fever (Cchf) in afghanistan: A teaching-case study | Africa | others | Health workforce | In-person training | Learning technique | Report |
| Al-Mayahi | 2019 | Surveillance gaps analysis and impact of the late detection of the first middle east respiratory syndrome case in south batinah, oman: A teaching case-study | Africa | others | Health workforce | In-person training | Learning technique | Report |
| Pelican | 2019 | Building an Ebola-Ready workforce: Lessons learned on strengthening the global workforce through university networks | International | Ebola | Health workforce | In-person training | Emergency plans | Report |
| Peters | 2019 | Development and pilot testing of an infection prevention and control (IPC) tool for humanitarian response to outbreaks and natural disasters | Zambia | others | Health workforce | In-person training | Emergency plans | Before-after |
| Keita | 2018 | Impact of infection prevention and control training on health facilities during the Ebola virus disease outbreak in Guinea | West Africa | Ebola | Health workforce | In-person training | Knowledge evaluation | Observational |
| Edem-Hotah | 2018 | Utilizing Nurses to Staff an Ebola Vaccine Clinical Trial in Sierra Leone during the Ebola Outbreak | Sierra Leone | Ebola | Health workforce | In-person training | Emergency plans | Report |
| Mbanjumucyo | 2018 | Major incident simulation in Rwanda: A report of two exercises | Rwanda | Disasters | Health workforce | Simulation | Emergency plans | Report |
| Bruning M.D | 2018 | The kid next door-raising awareness of Civilian health care providers to the needs of military children-a tactical approach to education | Iraq and Afganistan | War | Health workforce | In-person training | Knowledge evaluation | Before-after |
| Andrews R | 2018 | Computer-assisted disaster response: Benefits for global healthcare | Africa | Disasters | Health workforce | In-person training | Emergency plans | Report |
| Githuku | 2017 | Cholera outbreak in homa bay county, kenya, 2015 | Africa | Cholera | Health workforce | In-person training | Learning technique | Report |
| Jones-Konneh | 2017 | Intensive education of health care workers improves the outcome of ebola virus disease: Lessons learned from the 2014 outbreak in Sierra Leone | Sierra Leone | Ebola | Health workforce | In-person training- simulation | Impact | Report |
| Phaup | 2017 | Increasing access to HIV treatment and care services for key populations in zambia: A partnership approach to strengthening local capacity to provide sensitivity training to health workers | Zambia | others | Health workforce | In-person training | Emergency plans | Report |
| Nwandu | 2017 | Sustainable pepfar funded in service HIV training delivery models: A training impact evaluation from nigeria | Nigeria | others | Health workforce | In-person training | Knowledge evaluation | Before-after |
| Laudisoit | 2017 | A One Health team to improve Monkeypox virus outbreak response: An example from the Democratic Republic of the Congo | Democratic Republic of Congo | others | Health workforce | In-person training | Emergency plans | Report |
| Umar | 2017 | Learningthroughservice: 'shifa homes'a project of Shifa College of Medicine for rehabilitation of flood victims | Pakistan | Disasters | Citizens and affected population | In-person training | Emergency plans | Report |
| Vaz | 2016 | The role of the polio program infrastructure in response to Ebola virus disease outbreak in Nigeria 2014 | Nigeria | Ebola | National Institutions | In-person training | Emergency plans | Report |
| Houben | 2016 | TIME Impact - a new user-friendly tuberculosis (TB) model to inform TB policy decisions | International | others | National Institutions | Virtual training | Emergency plans | Report |
| Umoren | 2016 | From global to local: Virtual environments for global-public health education | International | Disasters | Citizens and affected population | Virtual training | Emergency plans | Report |
| Garde | 2016 | Implementation of the first dedicated Ebola screening and isolation for maternity patients in Sierra Leone | Sierra Leone | Ebola | Health workforce | In-person training | Impact | Report |
| Toda | 2016 | The impact of a SMS-based disease outbreak alert system (mSOS) in Kenya | Kenya | others | Health workforce | In-person training | Impact | RCT |
| Shao X | 2015 | Evaluation of anti-Ebola training system in the PLA Medical Team to Liberia and some suggestion | China | Ebola | Military | Blended | Emergency plans | Report |
| Ali | 2015 | Applicability of the advanced disaster medical response (ADMR) course, Trinidad and Tobago | Trinidad y Tobago | Disasters | Health workforce | In-person training | Learning technique | Before-after |
| Livingston | 2015 | Healthcare capacity building in Haiti: Training healthcare and non-healthcare providers in basic cardiopulmonary resuscitation | Haiti | Disasters | Health workforce and community | Simulation | Learning technique | Report |
| Berry | 2015 | How to set up an Ebola isolation unit: Lessons learned from Rokupa | Sierra Leone | Ebola | Health workforce | In-person training | Impact | Report |
| El-Bahnasawy | 2015 | Mosquito borne West Nile virus infection as a major threat | Egypt | others | Health workforce | In-person training | Impact | Report |
| Ariel | 2014 | The birth of family therapists: The kosova systemic family therapy training program | Kosovo | War | Health workforce | In-person training | Learning technique | Report |
| Reynolds | 2014 | Training health workers for enhanced monkeypox surveillance, Democratic Republic of the Congo | Congo | others | Health workforce | In-person training | Impact | Before-after |
| Acevedo | 2013 | Organization of the health system response to the 2009 H1N1 influenza pandemic in a hospital in Lima, Peru | Peru | others | Health workforce | In-person training | Emergency plans | Report |
| Hasanovic | 2013 | EMDR training for bosnia-herzegovina mental health workers in sarajevo as continuity of the building of psychotherapy capacity aftermath the 1992-1995 war | Bosnia-herzegovina | War | Health workforce | virtual training | Impact | Report |
| Hasanovic | 2013 | Training of bosnia-herzegovina mental health professionals in group analysis as the factor of development of culture of dialogue in the aftermath of the 1992-1995 war | Bosnia-herzegovina | War | Health workforce | In-person training | Impact | Report |
| Diaz | 2013 | Development of a severe influenza critical care curriculum and training materials for resource-limited settings | International | others | Health workforce | In-person training | Learning technique | Report |
| Asgary | 2013 | Comprehensive on-site medical and public health training for local medical practitioners in a refugee setting | Africa | Disasters | Health workforce | In-person training | Knowledge evaluation | Before-after |
| Plani | 2012 | Development of a hospital disaster plan and training exercises for chris hani baragwanath academic hospital and resource-limited countries | South Africa | Disasters | Health workforce | In-person training | Learning technique | Report |
| Wurapa | 2012 | Establishing a tropical medicine training program for the us department of defense (DOD) in kintampo, ghana: Overcoming challenges | Ghana | Disasters | Health workforce | In-person training | Emergency plans | Report |
| Chihanga | 2012 | Toward malaria elimination in Botswana: A pilot study to improve malaria diagnosis and surveillance using mobile technology | Botswana | others | Health workforce | virtual training | Learning technique | Report |
| Garnett | 2012 | Using south-south collaboration to strengthen midwifery skills and competencies in South Sudan | South Sudan | War | Health workforce | In-person training | Emergency plans | Report |
| Grosso | 2012 | The role of the international anaesthetist in the professional training and OT management of the anaesthesia nurses. 12 years of experience of emergency Italian NGO in Afghanistan | Afghanistan | War | Health workforce | In-person training | Emergency plans | Report |
| Norton | 2012 | The power of immersion; Training health personnel for disaster humanitarian responses | International | Disasters | Health workforce | In-person training | Emergency plans | Report |
| He | 2011 | The urgent rehabilitation technique education program for Wenchuan earthquake victims | China | Earthquake | Health workforce | In-person training | Emergency plans | Report |
| Sadiwa | 2011 | Addressing developmental delays among African children in post-conflict areas: An E-health approach | sierra Leone | War | Experts and volunteers | virtual training | Learning technique | Report |
| Oliveira | 2011 | Esperience in confronting the H1N1 epidemy | Brasil | others | Health workforce | In-person training | Impact | Report |
| Mbabazi | 2011 | Phase 1 implementation of male circumcision as a comprehensive package of HIV prevention in Rwanda | Rwanda | others | Health workforce | In-person training | Emergency plans | Report |
| Darby | 2011 | Multi-modal training for adult ICU nurses caring for paediatric patients in a war zone | Afghanistan | War | Health workforce | In-person training | Knowledge evaluation | Report |
| Way | 2011 | A modality of disaster response: Cyclone nargis and psychological first aid | Burma | Disasters | Health workforce and community | In-person training | Emergency plans | Report |
| Hasanovic | 2011 | Emdr training for mental health therapists in postwar bosniaherzegovina who work with psycho-traumatized population for increasing their psychotherapy capacities | Bosnia-herzegovina | War | Health workforce | In-person training | Emergency plans | Report |
| Oleribe | 2010 | From strategy to action: The vital roles of trained field epidemiologists and laboratory management professionals in epidemic control and prevention in Tanzania | Tanzania | others | Health workforce | In-person training | Emergency plans | Report |
| Kuhls | 2009 | International disaster training: Advanced disaster life support (ADLS) improves Thai physician and nurse confidence to respond to mass casualty disasters | Thailand | Disasters | Health workforce | In-person training | Knowledge evaluation | Before-after |
| van der Walt | 2006 | The effect of a CPD training (educational) intervention on the level of HIV knowledge of pharmacists | South Africa | others | Health workforce | virtual training | Knowledge evaluation | RCT |
| Peltzer | 2006 | A controlled study of an HIV/AIDS/STI/TB intervention with traditional healers in KwaZulu-Natal, South Africa | South Africa | others | Health workforce | In-person training | Knowledge evaluation | RCT |
| Wondmikun | 2005 | Successful coupling of community attachment of health science students with relief work for drought victims | Ethiopia | Disasters | Health workforce | In-person training | Impact | Report |
| Kabir | 2021 | Association between preference and e-learning readiness among the Bangladeshi female nursing students in the COVID-19 pandemic: a cross-sectional study | Bangladesh | COVID-19 | Health workforce | Virtual training | Learning technique | Cross-sectional |
| AlOsta | 2023 | Jordanian nursing students' engagement and satisfaction with e-learning during COVID-19 pandemic | Jordania | COVID-19 | Health workforce | Virtual training | Learning technique | Report |
| Severini | 2023 | How to incorporate telemedicine in medical residency: A Brazilian experience in pediatric emergency | Brasil | COVID-19 | Health workforce | in-person training | Learning technique | Before-after |
| Conyers | 2023 | Where There's a War, There's a Way: A Brief Report on Tactical Combat Casualty Care Training in a Multinational Environment | International | War | Military | In-person training | Emergency plans | Report |
| Farhat | 2022 | The educational outcomes of an online pilot workshop in CBRNe emergencies | Middle-east | Disasters | Health workforce | Virtual training | Emergency plans | Before-after |
| Mitchell | 2023 | Multimodal learning for emergency department triage implementation: experiences from Papua New Guinea during the COVID-19 pandemic | Papua New Guinea | Disasters | Health workforce | Virtual training | Knowledge evaluation | Before-after |
| Wang | 2022 | Rapid virtual training and field deployment for COVID-19 surveillance officers: experiences from Ethiopia | Ethiopia | COVID-19 | Health workforce | Virtual training | Emergency plans | Report |
| Suresh | 2021 | Predeployment training of Army medics assigned to prehospital settings | International | War | Health workforce | N/A | Knowledge evaluation | Report |
| Popova | 2022 | EXPERIENCE IN ORGANIZING URGENT TRAINING FOR GI PROFESSIONALS ON EMERGENCY CARE FOR ABDOMINAL INJURIES DURING THE WAR IN UKRAINE | Ukraine | War | Health workforce | Virtual training | Emergency plans | Report |
| Beckmann | 2022 | Training of psychotherapists in post-conflict regions: A Community case study in the Kurdistan Region of Iraq | Iraq | War | Health workforce | In-person training | Emergency plans | Report |
| Susanti | 2022 | The Effect of Caring Training on the Implementation of Caring Behavior and Work Culture of Nurses in Providing Services to COVID-19 Patients in an Indonesia's National Referral Hospital | Indonesia | COVID-19 | Health workforce | In-person training | Impact | Before-after |
| Canavese | 2022 | Massive Open Online Courses as Strategies to Address Violence through the Training of Health and the Intersectoral Professionals in Brazil | Brasil | COVID-19 | Health workforce | Virtual training | Learning technique | Report |
| Han | 2022 | Effect Analysis of "Four-Step" Training and Assessment Tool in the Prevention and Control of COVID-19 | China | COVID-19 | Health workforce | Virtual training | Learning technique | Report |
| Wood | 2022 | Evaluation of virtual online delivery of United Nations Office on Drugs and Crime (UNODC) national training on novel psychoactive substances (NPS) to healthcare professionals in Mauritius and the Seychelles during the COVID-19 pandemic | Mauritius and the Seychelles | COVID-19 | Health workforce | Virtual training | Knowledge evaluation | Before-after |
| Kamal | 2022 | Virtual Training on IGRT: A Unique Initiative of a Private Cancer Center from a Developing Country with Regional Academic Collaboration during COVID-19 Pandemic | Bangladesh | COVID-19 | Health workforce | Virtual training | Knowledge evaluation | Report |
| Fernandes Canesin | 2022 | Use of an innovative humanized virtual digital interactive heart failure clinical cases training strategy for cardiologist in the covid 19 pandemic | Brazil- Portugal and US | COVID-19 | Health workforce | Virtual training | Learning technique | Observational |
| Toro | 2022 | A Simulated Hospital in a COVID-19 Pandemic Environment for Undergraduate Neurology Students | Colombia | COVID-19 | Health workforce | simulation | Knowledge evaluation | Observational |
| Wong | 2022 | Better Surgical Ward Round: Replicating Near-Peer Teaching (NPT) on a virtual international platform during the COVID-19 pandemic organized by the IASSS | International | COVID-19 | Health workforce | Virtual training | Learning technique | Before-after |
| Ordonez Juarez | 2022 | Virtual Learning Environment for Surgery Residents in a Third Level Hospital at Mexico City, a Teaching Alternative | Mexico | COVID-19 | Health workforce | Virtual training | Knowledge evaluation | Report |
| Payne | 2022 | The Development and Evaluation of Online Home Palliative Training During COVID-19 Pandemic in South Africa | South Africa | COVID-19 | Health workforce | Virtual training | Impact | Report |
| Kumar | 2022 | Effectiveness of virtual versus inperson training of the FCCS course: A comparative study | Ghana, Nigeria, Augusta | COVID-19 | Health workforce | Virtual training | Knowledge evaluation | Observational |
| Kalayasiri | 2021 | Training of psychiatry and mental health in a low- and middle-income country: Experience from Thailand before and after COVID-19 outbreak | Thailand | COVID-19 | Health workforce | Virtual training | Emergency plans | Report |
| Tang | 2021 | Combat casualty care training of Chinese peacekeeping military doctors: An evaluation of effectiveness | China | War | Military | In-person training | Knowledge evaluation | Report |
| Siddiqui | 2021 | The impact of a "one day basic intensive care training program" on knowledge of non-intensivists during the COVID-19 pandemic | India | COVID-19 | Health workforce | In-person training | Knowledge evaluation | Before-after |
| Groninger | 2021 | Project echoTM palliative care: Impact of TELE-mentoring and teaching for healthcare providers working with rohingya refugees in Bangladesh | Bangladesh | War | Health workforce | Virtual training | Emergency plans | Report |
| Daniel | 2021 | Evaluation of the faculty experience in developing and delivering palliative care e-resource toolkit for COVID-19 for low and middle income countries (LMICS) | international | COVID-19 | Health workforce | Virtual training | Emergency plans | Before-after |
| Kharel | 2021 | Impact of a virtual COVID-19 trainer of trainers program implemented via an academic-humanitarian collaboration | international | COVID-19 | Health workforce | Virtual training | Learning technique | Before-after |
| Thakre | 2020 | Evaluation of effectiveness of Covid-19 training and assessment of anxiety among nurses of a tertiary health care center during the Corona Virus pandemic-an experimental study | India | COVID-19 | Health workforce | In-person training | Emergency plans | Report |
| Rishipathak | 2021 | Assessing the effectiveness of online teaching methodology among emergency medical professionals in Pune, India | India | COVID-19 | Health workforce | Virtual training | Knowledge evaluation | Report |
| Khoja | 2016 | Impact of simple conventional and Telehealth solutions on improving mental health in Afghanistan | Afghanistan | COVID-19 | Health workforce | Virtual training | Emergency plans | Report |
| Bernstein | 2022 | The Power of Connections: AAP COVID-19 ECHO Accelerates Responses During a Public Health Emergency | USA | COVID-19 | Health workforce | Virtual training | Learning technique | Report |
| Hunt | 2022 | Facilitating Real-Time, Multidirectional Learning for Clinicians in a Low-Evidence Pandemic Response | USA | COVID-19 | Health workforce | Virtual training | Impact | Report |
| Hunt | 2021 | Virtual Peer-to-Peer Learning to Enhance and Accelerate the Health System Response to COVID-19: The HHS ASPR Project ECHO COVID-19 Clinical Rounds Initiative | USA | COVID-19 | Health workforce | Virtual training | Impact | Report |
| Lingum | 2021 | Building Long-Term Care Staff Capacity During COVID-19 Through Just-in-Time Learning: Evaluation of a Modified ECHO Model | Canada | COVID-19 | Health workforce | Virtual training | Impact | Report |
| Stephens | 2022 | Adapting a Telehealth Network for Emergency COVID-19 Pandemic Response, 2020-2021 | International | COVID-19 | Health workforce | Virtual training | Impact | Report |
| Begay | 2021 | Strengthening Digital Health Technology Capacity in Navajo Communities to Help Counter the COVID-19 Pandemic | USA | COVID-19 | Health workforce | Virtual training | Emergency plans | Report |

Table 4: Type of study design

| *Study design* | N |
| --- | --- |
| Descriptive | 173 |
| Before-after | 87 |
| RCT | 18 |
| Cross-sectional | 14 |
| Observational | 12 |
| Review | 7 |
| Qualitative | 5 |
| Opinion | 3 |
| **Total** | **319** |

Table 5: Learner distribution across studies

| *Learner* | N |
| --- | --- |
| Health workforce | 268 |
| Military | 13 |
| Health workforce and community | 12 |
| Citizens and affected population | 12 |
| Experts and volunteers | 9 |
| National Institutions | 4 |
| Academia | 1 |
| **Total** | **319** |
